# Integration of clinical and genomic data defines prognostic phenotypes in resected perihilar cholangiocarcinoma: a national multicenter study

**DOI:** 10.64898/2026.02.16.26346384

**Authors:** Víctor López-López, Fernando Lucas-Ruiz, Cecilia Maina, Ana Isabel Antón-García, Laura Lladó, Marina Vila-Tura, Teresa Serrano, Rafael López-Ándujar, David Calatayud, Judith Perez-Rojas, José Ángel López-Baena, Isabel Peligros, Luis Sabater-Ortí, Isabel Mora-Oliver, Clara Alfaro-Cervello, David Pacheco, Enrique Asensio-Díaz, Beatriz Madrigal Rubiales, Cristina Dopazo, Concepción Gómez-Gavara, María Teresa Salcedo-Allende, Miguel Ángel Gómez-Bravo, Carmen Bernal Bellido, Juan José Borrero Martin, Alejandro Serrablo, Leyre Serrablo, Carlos Hörndler, Gerardo Blanco-Fernández, Isabel Jaén-Torrejimeno, Mario Díaz Delgado, Dilmurodjon Eshmuminov, Sergio Hernández-Kakauridze, Daniel Vidal-Correoso, Carlos Manuel Martínez-Cáceres, Jesús Miguel de la Peña-Moral, Pablo Ramírez, Alberto Baroja-Mazo, Ricardo Robles-Campos

## Abstract

**Background & Aims:** Perihilar cholangiocarcinoma is an aggressive malignancy with clinical heterogeneity and poor long-term outcomes after resection. Current prognostic assessment relies mainly on anatomical staging and pathological features, which incompletely capture the entire postoperative risk. We aimed to determine whether integrative analysis of clinical, surgical, pathological and tumor genomic data could improve time-resolved, individualized recurrence-risk prediction after curative-intent resection.

**Methods:** We performed a multicenter retrospective study including patients undergoing curative-intent resection for perihilar cholangiocarcinoma in ten Spanish hospitals (2003-2023). Overall and disease-free survival were analyzed using Cox models. Outcome-agnostic clinical phenotypes were derived by unsupervised clustering of clinical and surgical features. Targeted tumor sequencing of cancer-associated hotspot regions and selected genes was performed. Prognostic models integrating clinical and genomic data were trained and evaluated in independent training/test sets using penalized and latent-component Cox frameworks, with time dependent discrimination.

**Results:** The final cohort comprised 142 patients, with a median follow-up of 26.4 months. Recurrence occurred in 61.3% of patients, and 53.5% died during follow-up. Classical pathological factors were strongly associated with survival and recurrence. Unsupervised outcome-agnostic clustering identified three reproducible clinical phenotypes with markedly different recurrence patterns and survival, only partially explained by anatomical staging. Integrative clinical-genomic modelling further improved recurrence-risk prediction, achieving high discrimination in independent validation (time-dependent AUC ∼0.8). Moreover, the integrative model assigned higher risk over time to patients who relapsed. Patients combining unfavorable clinical phenotype with high genomic-derived risk exhibited a high probability of early recurrence.

**Conclusions:** Integrated clinical phenotyping and targeted genomic profiling substantially refine recurrence-risk stratification after resection of perihilar cholangiocarcinoma beyond anatomical staging alone. This provides a pragmatic framework for risk-adapted postoperative surveillance and therapeutic decision-making.

**Impact and Implications:** This study provides a data-driven framework integrating clinical, surgical and targeted genomic information to refine prognostic stratification after resection of perihilar cholangiocarcinoma, addressing the limitations of anatomy-based staging in capturing biological heterogeneity. The results are particularly relevant for clinicians managing postoperative surveillance and adjuvant strategies, as they identify patient subgroups with markedly different risks of early recurrence despite similar conventional staging. In practical terms, the combination of unsupervised clinical phenotyping and a targeted, biologically informed genomic panel could support risk-adapted follow-up intensity, selection for adjuvant or experimental therapies, and enrolment into clinical trials. While prospective validation is required before routine implementation, this approach offers a feasible and interpretable pathway toward precision postoperative management in a highly aggressive malignancy.

## Introduction

Perihilar cholangiocarcinoma (pCCA) is the most common and one of the most aggressive malignancies of the biliary tract, exhibiting marked heterogeneity in its clinical behavior. Surgical resection remains the only potentially curative treatment; however, only a minority of patients are eligible for radical surgery, and recurrence rates remain high even after complete resection. Long-term outcomes are therefore poor, with five year survival rates rarely exceeding 40%, reflecting the intrinsic biological aggressiveness of the disease [2].

Current prognostic assessment in pCCA relies primarily on anatomical staging systems and histopathological parameters, including tumor stage, lymph node involvement, resection margin status, and differentiation grade [3]. While these factors are clinically informative, they fail to fully capture the biological complexity and heterogeneity of the disease, resulting in substantial variability in outcomes among patients with similar staging profiles [4]. This limitation highlights the need for integrated prognostic approaches that extend beyond conventional staging and incorporate the multifactorial determinants of tumor behavior. Advances in cancer genomics have substantially enhanced the understanding of tumor biology, revealing that patients with apparently similar clinicopathological characteristics may experience markedly different oncological outcomes [5]. This paradigm shift has highlighted the limitations of traditional prognostic models and has underscored the role of molecular heterogeneity as a major determinant of disease behavior and clinical evolution.

In this clinical context, systemic therapies have shown modest benefits in advanced biliary tract cancers, yet their impact in the postoperative and recurrence settings remains limited and difficult to predict [6]. As a consequence, there is a clear clinical need for tools capable of more accurately identifying patients at high risk of early recurrence and poor survival, as well as those with a more favorable prognosis who may benefit from standard surveillance strategies and avoid ineffective overtreatment.

Recent molecular studies have identified recurrent genetic alterations in biliary tract cancers, including mutations affecting TP53, CDKN2A/2B, ARID1A, KRAS, and chromatin-remodeling pathways, which have been associated with tumor aggressiveness, early recurrence, and resistance to therapy [7]. Despite these advances, the clinical translation of molecular findings has been hampered by the complexity of genomic data and by the limited prognostic value of isolated biomarkers when considered outside their clinical and pathological context [8]. Importantly, genomic alterations do not act in isolation but interact with clinical variables, tumor microenvironment, and surgical factors to shape disease evolution [9, 10].

To address this challenge, integrative analytical strategies that combine tumor genomic profiles with key clinical, surgical and pathological variables have emerged as a promising approach. Unsupervised clustering techniques enable the identification of clinically meaningful patient subgroups based on shared multidimensional characteristics, while modern survival modelling frameworks allow the incorporation of high-dimensional genomic data alongside clinical information to generate robust, interpretable prognostic signatures [11, 12]. These methodologies provide a holistic assessment of risk that better reflects the multifactorial nature of oncological outcomes.

The present study was therefore designed to evaluate whether the joint analysis of clinical, surgical, pathological, and genomic features, using state-of-the-art data-driven analytical methods, can improve prognostic stratification in patients undergoing curative-intent resection for pCCA. By integrating routinely collected clinical data with targeted tumor genomic profiling, we aimed to identify distinct prognostic groups with significantly different risks of recurrence and survival, thereby advancing precision prognostication in this highly aggressive malignancy.

## Patients and Methods

### Study design and patient cohort

We conducted a multicenter retrospective study including patients with pCCA who underwent curative-intent surgical resection between February 2003 and February 2023 across ten Spanish referral centers (Fig. 1A). Inclusion and exclusion criteria are detailed in the Supplementary Methods. The study protocol was approved by the central ethics committee of Hospital Universitario Virgen de la Arrixaca (Murcia, Spain; registry number 2021-4-7-HCUVA) and by the institutional review boards of all participating centers.

**Fig. 1.** Study cohort and outcome distributions. (A) Schematic overview of patient inclusion criteria and downstream clinical and genomic analyses across 10 Spanish hospitals (2003-2023). (B-C) Distribution of days to last follow-up/exitus (B) or to recurrence (C), displayed as density plot with overlaid boxplot and individual patient values. (D-E) Pie chart of overall survival categories (<1 year, 1-3 years, >3 years) (D), exitus (E) or recurrence (F) status.

Baseline demographic, clinical, laboratory, surgical, and pathological variables were systematically collected. Overall survival (OS) was defined as the time from surgery to death from any cause or last follow-up, and disease-free survival (DFS) as the time from surgery to first documented recurrence, with censoring at last follow-up. Detailed variable definitions and follow-up procedures are provided in the Supplementary Methods.

### Statistical analysis of clinical outcomes

Associations between individual clinicopathological variables and OS or DFS were initially explored using univariable Cox proportional hazards models. Variables reaching nominal significance were further visualized using Kaplan-Meier survival curves. Comparative analyses across survival categories, recurrence status, and patient subgroups were performed using appropriate parametric or non-parametric tests depending on variable type and distribution. Full details of the statistical procedures, model specifications, and multiple-testing corrections are provided in the Supplementary Methods.

### Unsupervised clinical clustering

To identify prognostically relevant patient phenotypes in an outcome-agnostic manner, we performed unsupervised clustering using preoperative, intraoperative, and pathological variables. Mixed-type data were integrated using a Gower dissimilarity matrix, followed by divisive hierarchical clustering. Cluster stability and separation were assessed using silhouette analysis and low-dimensional visualization techniques. The prognostic relevance of the identified clusters was evaluated using Cox models and Kaplan-Meier analyses. The complete clustering workflow is described in the Supplementary Methods.

### Tumor sampling and targeted genomic profiling

Formalin-fixed paraffin-embedded tumor samples were retrieved for DNA extraction and genomic analysis. Targeted sequencing was performed using a custom amplicon-based panel covering cancer-associated hotspot regions and selected coding exons of ARID1A, ERBB3 and CDKN2B genes, with established relevance in biliary tract cancer biology. Sequencing, variant calling, annotation, and quality control procedures are detailed in the Supplementary Methods.

### Integrative clinical-genomic modelling

Integrative prognostic models were developed using DFS as the primary endpoint, combining clinical variables and genomic features within penalized and latent-component Cox modelling frameworks. Model development, hyperparameter optimization, internal cross-validation, and independent test-set evaluation were performed using the COXMOS platform [13]. Model performance was assessed using time-dependent AUC, concordance index, and calibration metrics. Methods for risk stratification, component interpretation, and proportional hazards assessment are described in detail in the Supplementary Methods.

### Statistical environment

All analyses were conducted in R (version 4.4.0). A full list of packages and software versions is provided in the Supplementary Methods.

## Results

### Clinical and demographic characteristics of the cohort

The study enrolled 192 patients from 10 Spanish centers from February 2003 to February 2023. After exclusion criteria, the final cohort comprised 142 patients undergoing hepatic resection with curative intent (Fig. 1A). The median follow-up (or time to death) was 26.4 months (804 days; IQR 1031.3) (Fig. 1B; Table 1). Among patients with recurrence, the median time to recurrence was 14.29 months (435 days; IQR 644.5) (Fig. 1C; Table 1). 23.9% of patients had <1 year of follow-up, 39.4% had 1-3 years, and 36.6% were followed for >3 years (Fig. 1D; Table 1). Finally, 76 patients (53.5%) had died and 87 (61.3%) had developed tumor recurrence during the follow-up (Fig. 1E-F; Table 1).

The median age at surgery was 67 years, with a slight male predominance. Preoperative assessment revealed frequent cholestasis, variability in CA 19-9 values, and a high rate of biliary drainage before surgery. Tumors were locally advanced (predominantly T2), with AJCC stage II nearly half of the cases. Nodal involvement was present in roughly one-quarter of patients. Histologically, most tumors were moderately differentiated; lymphovascular and perineural invasion were frequent. Microscopically positive resection margins (R1) were reported in about one-third (Table 1).

Major hepatectomies predominated, often requiring caudate lobe resection, vascular reconstruction or use of the Pringle manoeuvre. Postoperative morbidity was substantial, with major complications occurring in ∼40% of patients. Post-hepatectomy liver failure was uncommon but clinically relevant. Most patients received adjuvant chemotherapy, and around one-fifth received adjuvant radiotherapy (Table 1).

In summary, these data define a large, heterogeneous and clinically representative national cohort of resected pCCA, providing a robust foundation to explore prognostic drivers of survival, recurrence patterns and risk stratification beyond conventional staging.

### Tumor stage and invasion patterns drive OS and probability of recurrence

Variables showing significant prognostic value in univariable Cox proportional hazards analyses were explored using Kaplan-Meier curves for both survival and recurrence outcomes.

In OS analyses, node-positive disease (pN1), advanced AJCC stage (IIIA-IIIC), and perineural or vascular invasion identified subgroups with inferior OS. Preoperative biliary drainage with endoscopic bile duct stents or nasobiliary drainage (category B, EBS/ENBD) and a more advanced primary tumor T category (dichotomised at T2A) were associated with reduced OS (log-rank p < 0.05) (Fig. 2A).

**Fig. 2.** Kaplan-Meier survival analyses for clinicopathological variables. (A) OS curves showing survival probability on the y-axis. (B) DFS curves showing recurrence probability on the y-axis. Log-rank p-values are reported in each panel.

DFS curves showed a similar pattern. Node-positive status, perineural and vascular invasion, higher AJCC 8th edition stage, portal vein embolization (PVE) and preoperative percutaneous trans-hepatic biliary drainage (category C, PTCD) were linked to worse DFS. Surgical margin status also had a significant impact since patients with R1 resections experienced higher recurrence rates than those with negative margins. DFS further differed according to the administration of adjuvant chemotherapy (Fig. 2B).

In addition to these time-to-event analyses, we compared baseline characteristics across three OS categories (<1 year, 1-3 years and >3 years). In these classical comparisons, the presence of relevant comorbidities, advanced AJCC stage, node-positive disease, distal duodenal margin involvement, and recurrence status at last follow-up were significantly associated with worse OS (Table S1). Likewise, the factors significantly associated with shorter DFS included node-positive status, primary tumor stage, perineural invasion, higher AJCC stage, preoperative PTCD, surgical margin status and the administration of adjuvant chemotherapy (Table S2).

Overall, these analyses confirm that classical markers of tumor burden and invasive behavior, including nodal involvement, advanced stage, perineural and vascular invasion, and margin status, remain the dominant determinants of outcome, validating robustness of the cohort.

### Agnostic clustering delineates three clinically distinct risk profiles with divergent survival and recurrence

To identify clinically patient subgroups with distinct prognoses, we conducted an unsupervised clustering analysis designed to be fully agnostic by restricting inputs to preoperative and intraoperative variables, and excluding any information related to survival, recurrence or follow-up. A Gower-distance dissimilarity matrix was computed to integrate numerical, binary, and categorical variables. This matrix fed DIANA, a divisive hierarchical clustering algorithm without assumptions of continuous variables or spherical cluster structure.

Dendrogram and the corresponding silhouette plot supported a three-cluster solution as a reasonable compromise between parsimony and separation (Fig. 3A-B). Nearly all patients exhibited positive silhouette values, indicating cluster assignments were consistent with the underlying dissimilarity structure (Fig. 3B). t-SNE dimensionality reduction showed that the clinical information was sufficient to separate patients into the same three clusters in low-dimensional space (Fig. 3C).

**Fig. 3.** Unsupervised patient stratification by DIANA-Gower clustering and clinical annotation. (A) Hierarchical clustering dendrogram of the DIANA-Gower dissimilarity matrix, highlighting three patient clusters. (B) Silhouette plot showing cluster cohesion/separation for the three-cluster solution. (C) Two-dimensional t-SNE representation of patients colored by cluster assignment. (D-G) Kaplan-Meier curves stratified by cluster for OS (D), DFS shown as recurrence probability over 60 months (E), early OS (0-8 months) (F), and DFS probability (0-8 months) (G), with pairwise log-rank p-values. (H) t-SNE embedding annotated with key clinicopathological variables. For binary categorical variables, labels indicate the percentage of positive cases within each cluster, whereas for continuous variables, labels report the median and interquartile range. Cluster boundaries are overlaid.

Kaplan-Meier curves for OS up to 60 months of follow-up showed that clusters 1 and 2 had similar survival, whereas cluster 3 displayed worse outcomes. Pairwise comparisons confirmed this pattern showing difference between cluster 1 vs 3 (p = 0.014), while differences between cluster 2 vs 3 (p = 0.10) and cluster 1 vs 2 (p = 0.65) were not statistically significant (Fig. 3D). DFS analyses revealed a complementary pattern. Both clusters 1 and 2 showed lower recurrence risk with later and less frequent recurrences than cluster 3, with significant adjusted differences for cluster 1 versus 3 (p = 0.0019) and cluster 2 versus 3 (p = 0.032), while clusters 1 and 2 were broadly similar to each other (Fig. 3E).

An early-outcome analysis restricted to the first 8 months after surgery, showed the same trend for OS: Clusters 1 and 2 maintained higher early survival than cluster 3, although the pairwise differences did not reach conventional statistical significance (Fig. 3F). In the DFS analysis, cluster 3 again showed the highest recurrence rate, with a significant excess of events compared with cluster 1 (p = 0.0067), while cluster 2 occupied an intermediate position with a trend towards more recurrences than cluster 1 (p = 0.089) (Fig. 3G).

To characterize these agnostic clusters in more detail, we compared the distribution of clinical, surgical and pathological variables across groups (Fig. S1, Table S3-4) and projected them onto the t-SNE space (Fig. 3H). Cluster 3 concentrated adverse features, including higher preoperative bilirubin, more advanced T and AJCC stages, nodal involvement, advanced differentiated tumors, and higher rates of R1 resection together with lymphatic, vascular and perineural invasion (Fig. 3H, Fig. S1, Table S3-4).

Clusters 1 and 2 showed more favorable profiles, but with a clear gradient between them. Cluster 1 included patients with lower AJCC stage, negative nodes and lower rates of lymphovascular/perineural invasion and R1 margins, whereas cluster 2 showed an intermediate profile, with more comorbidities and more frequent use of complex procedures, including bile duct/left hepatectomy, segment-1 resection, Pringle manoeuvre, and perioperative transfusion, while its pathological features were less adverse than those observed in cluster 3 (Fig. 3H, Fig. S1, Table S3-4). When restricting the analysis to patients with ≤8 months of follow-up, differences in AJCC stage, nodal status and vascular invasion continued to separate cluster 3 from the others, while segment-1 resection remained characteristic of cluster 2 (Fig. S2, Table S5-6).

In summary, agnostic DIANA-Gower clustering delineates three distinct risk phenotypes that were not captured by anatomical staging alone. Cluster 1 represented a favorable profile with low recurrence and prolonged survival. Cluster 3 defined an aggressive phenotype marked by early recurrence and poor outcomes. Cluster 2 showed an intermediate pattern, with frequent recurrence but relatively preserved survival. Together, these findings highlight the added prognostic value of multifactorial clinical stratification.

### Integrative modelling identifies a sPLS-DACOX clinical-genomic signature for recurrence risk

In 116 patients with targeted genomic profiling, 49% of the 13,184 variants detected were synonymous, 37% of unknown functional consequence and 12% missense; nonsense and stop-loss variants were rare (0.8% and 0.007%, respectively) (Fig. 4A). The functional categories were distributed across the three genes with broadly similar proportions, although ARID1A and ERBB3 harbored the largest number of variants, as expected given that their coding sequences were specifically enriched in the panel (Fig. 4B).

**Fig. 4.** Genomic variant annotation and predictive modelling workflow. (A) Pie chart summarizing variant consequence classes. (B) Stacked bar plot showing the log2 number of altered genomic positions per gene, stratified by variant consequence classes. (C) Schematic of the modelling pipeline. (D) Left: time-dependent AUC trajectories across follow-up (months) for the evaluated methods. Right: boxplots summarizing model performance metrics across resampling iterations.

When we restricted the analysis to variants present in at least 10% of patients, we did not identify any single alteration that clearly distinguished recurrent from non-recurrent cases (Fig. S3), nor patients who died within 1 or 3 years from those with longer survival (Fig. S4). This suggested that no individual variant was sufficient to define recurrence risk, and that prognostic information is likely to emerge from the combined effect of multiple genetic changes together with clinical factors. We therefore used the COXMOS framework to build integrative survival models. The number of predictors greatly exceeded the number of events (EPV = 0.005), making classical Cox models unreliable and justifying the use of penalized or dimension-reduction methods.

For DFS, data were split into training (70%) and test (30%) sets with similar event rates (train data: 49 events/33 censored; test data: 20/14). We compared four modelling approaches: Elastic-Net, sPLS-ICOX, sPLS-DACOX and sPLS-DRCOX, using repeated 10-fold cross-validation and 10 repeats for each model, up to 8 components, adjusting the hyperparameters to optimize time-dependent AUC, C-index, or a combination of C-index and I.Brier criterion, and evaluating performance on the independent test set (Fig. 4C).

All penalized approaches showed acceptable discrimination in the independent test set, with time-dependent AUC values generally ranging from 0.6 to 0.9 across follow-up. However, overall performance differed across methods, with sPLS-DACOX providing the most consistently high discrimination over time (Fig. 4D).The sPLS-DACOX model also achieved an optimal integrated Brier Score, C-index and AIC (Fig. S5A-C). By contrast, the sPLS-DRCOX model attained the highest C-index in the test set (0.88), indicating superior concordance between predicted and observed survival times (Fig. 4D). Therefore, both models with an optimal C-index were selected to investigate in greater detail the contribution of SNPs and clinical variables to survival.

We checked the proportional hazards assumption using Schoenfeld tests. These tests showed no evidence of violations (global p = 0.809 for sPLS-DACOX; p = 0.357 for sPLS-DRCOX), with non-significant component-wise tests and residuals fluctuating around zero without time-dependent patterns (Fig. S6-7). This supports the validity of both models for time-to-event prediction in our cohort.

To visualize model behavior, we examined sPLS-DACOX linear predictor (LP) distributions stratified by recurrence status. Patients who recurred showed a clear rightward shift towards higher LP values, with limited overlap (Fig. 5A). Projection onto the first two latent components confirmed partial separation between groups (Fig. 5B). The sPLS-DRCOX model showed a similar but more modest trend, with greater overlap in the component-score space (Fig. S8A-B), and was therefore not prioritized for subsequent downstream analyses.

**Fig. 5.** sPLS-DACOX recurrence model outputs and feature contributions. (A) Density distribution of the model-derived LP stratified by event status. (B) Sample projection on the first two latent components, colored by recurrence status. (C) Forest plot of HR and 95% confidence intervals for the latent components, with associated p-values. (D) Kaplan-Meier curve of recurrence probability from 0 to 60 months, stratified by LP-based risk groups using a cut-off optimized to maximize separation between groups, with log-rank p-value. (E-F) Individual predicted recurrence probability trajectories every 2 months up to 60 months, colored by event status (E) or DIANA-Gower cluster assignment (F). (G) Bar plot of the top 50 variables ranked by absolute model coefficient (β), indicating their direction and contribution to the sPLS-DACOX model. (H) Bar plot of the genes mapped from the selected variants, showing the number of model-selected variants per corresponding gene.

Components 1 to 7 were significantly associated with recurrence risk and showed hazard ratios with relatively narrow confidence intervals with a statistical significance (Fig. 5C), whereas component 8 was not. These findings suggest that the prognostic signal is distributed across several orthogonal components, capturing complementary patterns of clinical and genetic variation.

Time-dependent AUC analyses supported the multicomponent nature of sPLS-DACOX. In the training set, the full LP achieved the highest discrimination by integrating all latent components. (Fig. S9A). In the independent test set, the full LP remained the best overall summary, with component 1 only slightly outperforming it at very late time points when few patients were at risk (Fig. S9B), suggesting that prediction relies on complementary signals across components.

The optimal LP threshold was -0.77 for sPLS-DACOX model, yielding significant separation in the training cohort (log-rank p <0.001). Applied to the test set, the threshold showed a trend towards poorer outcomes in the high-risk group (p = 0.068; Fig. S9C) and became significant when restricting to follow-up < 60 months (p = 0.02; Fig. 5D). At individual level, predicted recurrence probabilities every two months were consistently higher in patients who relapsed, confirming effective risk assignment (Fig. 5E).

Overlaying sPLS-DACOX-predicted recurrence probabilities and trajectories by DIANA-Gower cluster, revealed a clear pattern (Fig. 5F). Cluster 3 concentrated the steepest recurrence curves, while cluster 1 and 2 aggregated the lowest-risk trajectories. This shows that sPLS-DACOX model not only predicts individual recurrence risk but also recovers the latent risk structure revealed by the unsupervised clustering, thereby demonstrating concordance between the two approaches.

Pseudo-beta survival weights of the variables contributing to the dynamic components, showed that 84 variables explained 100% of the sPLS-DACOX model, with the top 50 accounting for 88.61% (Fig. 5G, Table S7). Clinical covariates featured prominently among the highest-weighted predictors, including comorbidity burden, vascular invasion, grading, post-hepatectomy liver failure, adjuvant radiotherapy, PVE, blood transfusion, hospital readmission, surgical technique, and number of lymph nodes resected (Fig. 5G, Table S7).

Likewise, the genomic variants selected mapped to 19 cancer-related genes. ARID1A contributed the highest number, followed by CDKN2A, EGFR and FLT3, with additional variants involved in cell-cycle regulation (CDKN2A/B, TP53, APC), chromatin remodeling (ARID1A, SMARCB1) and RTK/PI3K–AKT signaling (EGFR, ERBB3/4, KIT, FLT3, KDR, PIK3CA, AKT1, RET) (Fig. 5H).

Models for OS showed signs of overfitting (Fig. S10A-B), such as the first latent component outperforming the full LP in test data; therefore, these were discarded.

Taken together, these results show that recurrence risk is effectively captured by an integrative clinical-genomic signature where multiple clinical variables and variants jointly define risk trajectories with robust performance.

## Discussion

In this national multicenter cohort of resected pCCA, we demonstrate that postoperative outcome is driven not only by classical pathological features but by the multifactorial convergence of clinical phenotype, surgical complexity and tumor genomics, which together define biologically distinct disease trajectories. Through agnostic clinical clustering and integrative clinicogenomic modelling, we identified reproducible high-, intermediate- and low-risk groups with markedly different patterns of recurrence and survival that are only partially captured by conventional AJCC staging. These findings support the concept that pCCA should not be viewed as a single prognostic entity after resection, but rather as a spectrum of biologically defined subtypes that can be recognized using routinely available clinical, surgical and pathological variables complemented by targeted genomic profiling.

Our overall survival estimates (median ≈40 months; ∼40% at 5 years) are consistent with contemporary surgical series [14–19], and perioperative morbidity and mortality remain substantial even in experienced centers [19]. Together, these observations underscore that, despite technical and perioperative advances, tumor biology remains the dominant driver of postoperative outcomes in pCCA, as reflected by the high frequency of recurrence following curative-intent resection and the persistent need for more effective adjuvant and systemic strategies.

Consistent with prior evidence [20–25], classical pathological factors such as lymph node involvement, positive resection margins and poor tumor differentiation remained strong determinants of overall and disease-free survival in our cohort. These established features continue to provide essential prognostic information, but their limited ability to fully account for the observed heterogeneity in recurrence patterns and survival highlights the need for integrative approaches that extend beyond conventional clinicopathological assessment.

Beyond individual pathological risk factors, outcome-agnostic clinical clustering identified three reproducible phenotypes that reflect distinct postoperative trajectories not fully captured by AJCC/TNM staging. These phenotypes integrate tumor extent, biological aggressiveness, comorbidity burden and surgical complexity into coherent risk profiles with markedly different risks of recurrence and survival. Similar to prior work in cholangiocarcinoma describing biologically and clinically distinct subgroups that transcend anatomical classifications [26–28], our analysis reveals that patients undergoing resection for pCCA do not constitute a homogeneous prognostic population. In this framework, one phenotype was characterized by limited tumor burden and favorable pathological features and showed sustained postoperative survival, whereas another concentrated patients with advanced, node-positive disease and early systemic relapse. An intermediate phenotype displayed mixed clinical and pathological features, with high recurrence rates but relatively preserved overall survival. Together, these findings indicate that routinely collected perioperative variables can be leveraged to define clinically meaningful risk strata beyond anatomical stage alone.

The recurrence patterns associated with these phenotypes further support their biological plausibility. Patients within the high-risk phenotype showed a predominance of early, often multifocal systemic relapse, frequently occurring within the first postoperative year, a pattern consistently associated with aggressive tumor biology and limited long-term survival in prior series [25, 29, 30]. In contrast, the favorable-risk phenotype exhibited both lower recurrence rates and a temporal shift toward later relapse, suggesting a fundamentally different disease behavior, potentially related to a reduced invasive features and greater tumor differentiation. Importantly, these clinically defined phenotypes were derived exclusively from routinely available preoperative and intraoperative variables, in contrast to transcriptomic-based classifications [31], which remain costly, technically demanding and difficult to implement at scale. This pragmatic approach enhances translatability and supports the use of unsupervised clinical phenotyping as a tool for risk-adapted surveillance strategies.

Collectively, these results suggest that a substantial proportion of postoperative risk in pCCA can already be captured through integrative clinical phenotyping, without recourse to molecular data. This provides a clinically intuitive framework for early risk stratification.

Given the growing evidence that specific genetic alterations modulate prognosis and treatment response in biliary tract cancers [32–35], we deliberately adopted a targeted and biologically informed genomic strategy rather than broad untargeted profiling. By focusing on a limited panel of cancer-associated hotspots and coding regions in genes with established roles in chromatin remodeling, cell-cycle regulation and receptor signaling, we aimed to maximize biological relevance while preserving clinical feasibility in archival cohorts.

Integrating this restricted genomic panel with routinely available clinical and pathological variables resulted in a marked improvement in recurrence-risk prediction compared with clinical data alone. The resulting clinicogenomic model achieved high discrimination in independent validation, indicating that a parsimonious set of genomic features can add meaningful prognostic value without introducing noise or excessive model complexity. Importantly, risk prediction did not depend on a single dominant alteration but emerged from the combined contribution of multiple clinical and molecular factors, supporting the robustness and potential generalizability of the approach.

From a clinical perspective, the integration of genomic information refined risk stratification beyond anatomical staging and unsupervised clinical clustering alone. Patients with unfavorable clinical phenotypes who also exhibited high model-derived LP values represented a subgroup with an extreme risk of early systemic recurrence, a pattern that was consistently observed across analyses. Conversely, patients with favorable clinical profiles and low predicted genomic risk showed delayed or absent recurrence, even within the same AJCC stages. The genomic features selected by the model are biologically coherent with prior knowledge. Alterations involving TP53, CDKN2A/2B and ARID1A have repeatedly been linked to aggressive tumor behavior, genomic instability and poor outcomes in biliary tract cancers [32–34, 36]. Their co-selection alongside classical pathological variables underscores that tumor genotype and clinical phenotype jointly shape recurrence propensity. The presence of these variants, and in some cases their combinatorial patterns, within the prognostic signature suggests that the model is not merely capturing statistical correlations but reflects core biological processes driving early relapse. This reinforces the value of clinically deployable targeted panels focused on pathways with well-established functional impact.

Taken together, these findings indicate that the combination of unfavorable clinical phenotype and high genomic-derived risk identifies a subset of patients with particularly aggressive disease biology, who are unlikely to be adequately managed by standard postoperative strategies.

From a clinical standpoint, our findings have several immediate and pragmatic implications. First, the identification of reproducible clinical phenotypes through unsupervised clustering suggests that postoperative risk stratification can be substantially refined using variables that are already routinely collected in daily practice. Patients belonging to the high-risk clinical phenotype, particularly those characterized by advanced local disease, nodal involvement and adverse invasion patterns, represent a subgroup in whom early systemic relapse is highly probable and in whom standard surveillance strategies may be insufficient.

Second, the integration of targeted genomic information further sharpens this risk assessment. Patients with a high model-derived LP constitute a subgroup with extreme risk of early recurrence, often within the first postoperative year. These patients may be prioritized for intensified postoperative surveillance, earlier imaging schedules, and consideration of adjuvant or experimental systemic therapies, even when conventional staging alone might suggest intermediate risk. Conversely, patients with low predicted genomic risk may be candidates for standard follow-up strategies, potentially avoiding overtreatment and unnecessary toxicity.

Third, the use of a limited, biologically informed genomic panel enhances feasibility and interpretability, facilitating future clinical implementation. Unlike broad genomic profiling approaches that are costly and difficult to integrate into routine workflows, a restricted panel focused on key pathways associated with tumor aggressiveness offers a realistic path towards decision-support tools, such as postoperative risk calculators or stratification algorithms. These tools could assist multidisciplinary teams in tailoring surveillance intensity, selecting patients for adjuvant trials, and designing genotype-informed translational studies.

Collectively, our results support a shift towards integrated clinical-genomic decision-making in resected pCCA, where postoperative management is guided not solely by anatomical stage, but by a composite assessment of tumor biology, clinical phenotype and genomic risk.

We acknowledge several limitations. The targeted genomic panel used in this study, while pragmatic and biologically informed, does not capture the full spectrum of tumor heterogeneity that could be accessed through whole-exome approaches. Although the integrative model demonstrated robust performance in an independent test set, the multicenter and retrospective nature of this 20-year cohort highlights the need for broader external validation before clinical implementation. Finally, while the identified clinical-genomic signatures are biologically plausible, functional validation of selected variants and integration with transcriptomic or tumor microenvironmental data would be necessary to strengthen causal interpretation.

## Conclusions

Overall, this national multicenter study shows that prognosis after curative-intent resection for pCCA is best understood as the result of convergent, multifactorial determinants rather than anatomical stage alone. Our results support a precision-medicine framework in which postoperative surveillance and therapeutic strategies are informed by integrated clinical and genomic risk profiles. Prospective validation and extension to additional molecular layers will be essential to determine whether such stratification can ultimately modify clinical decision-making and improve outcomes in high-risk patients.

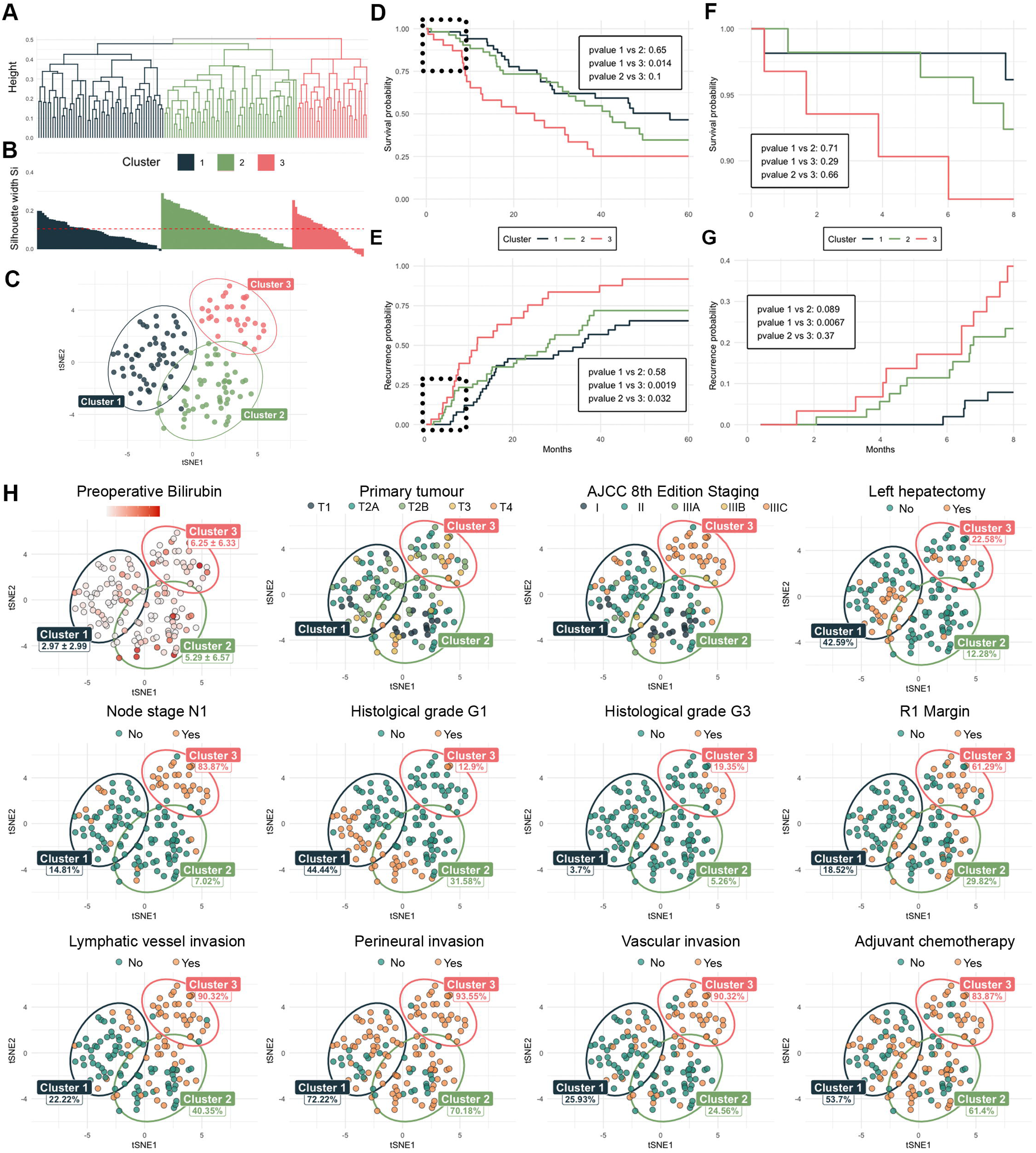

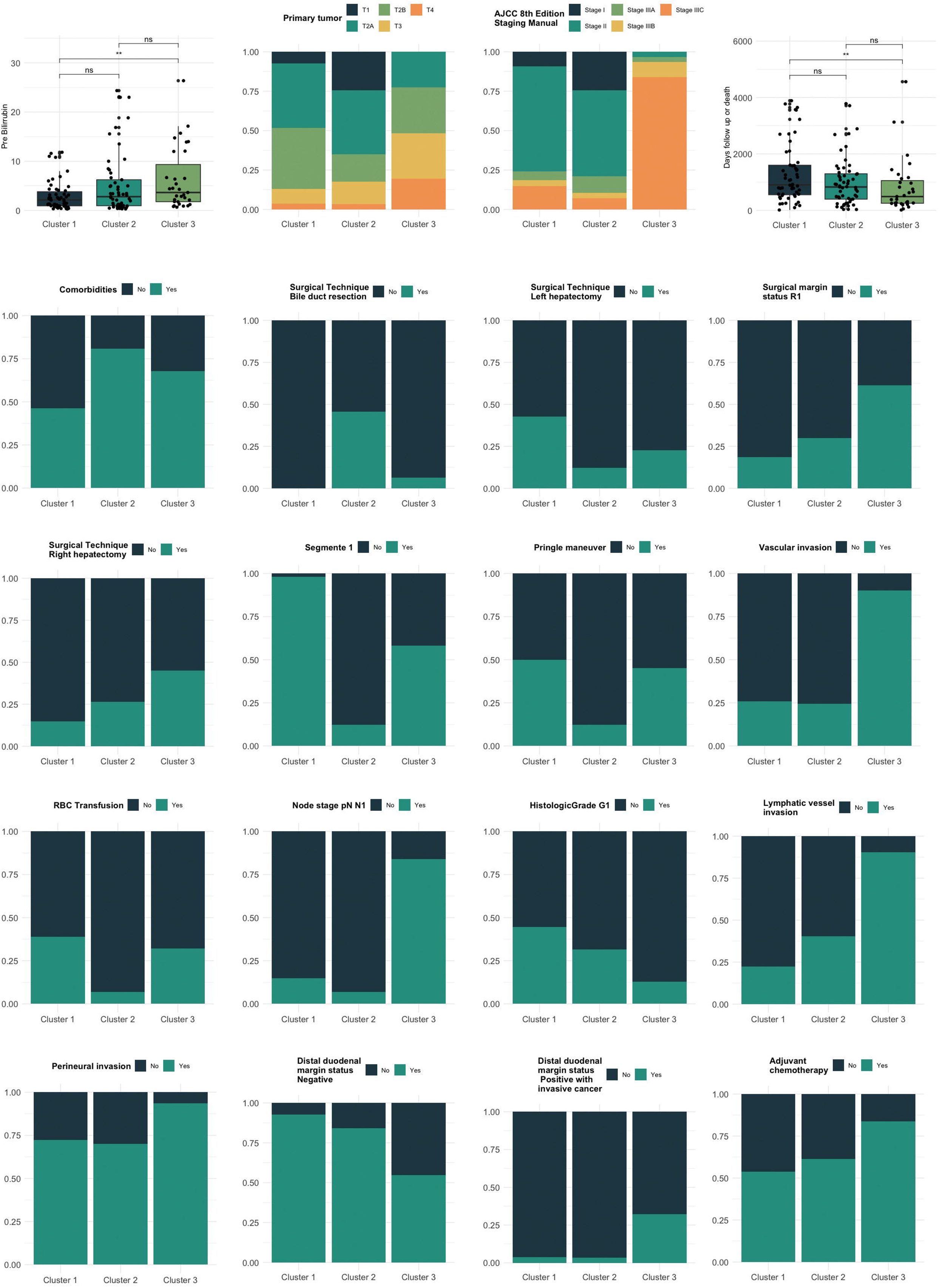

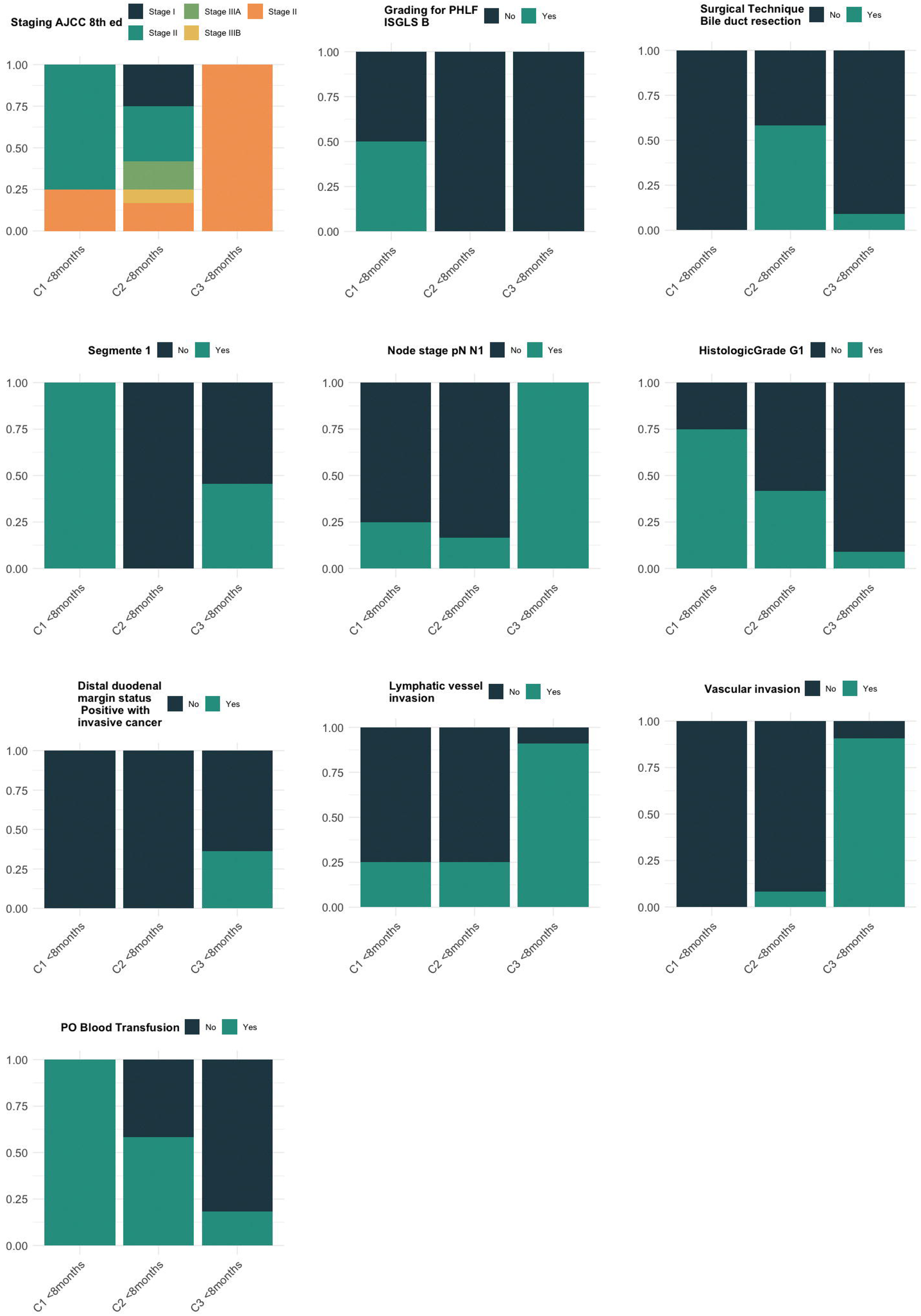

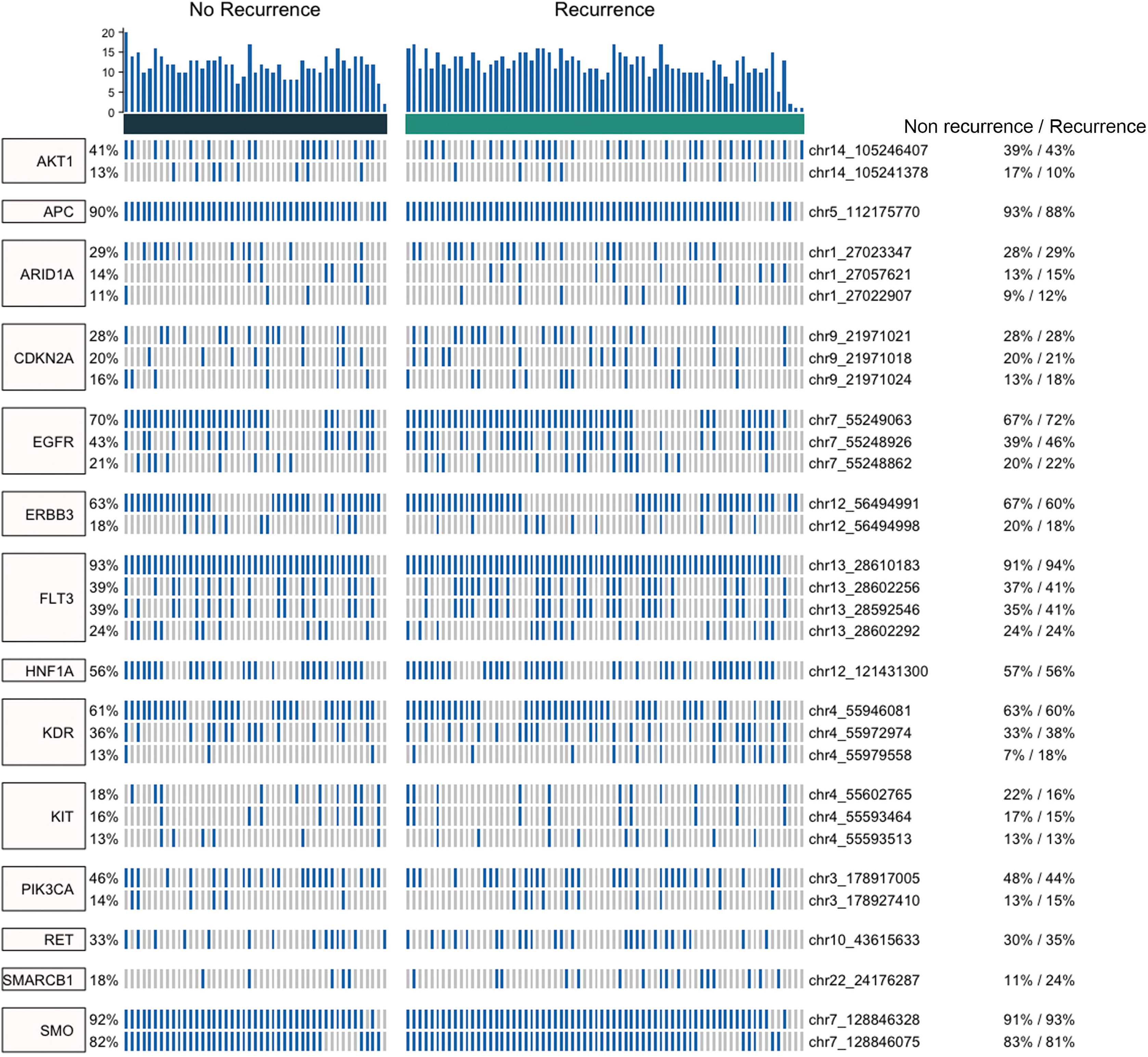

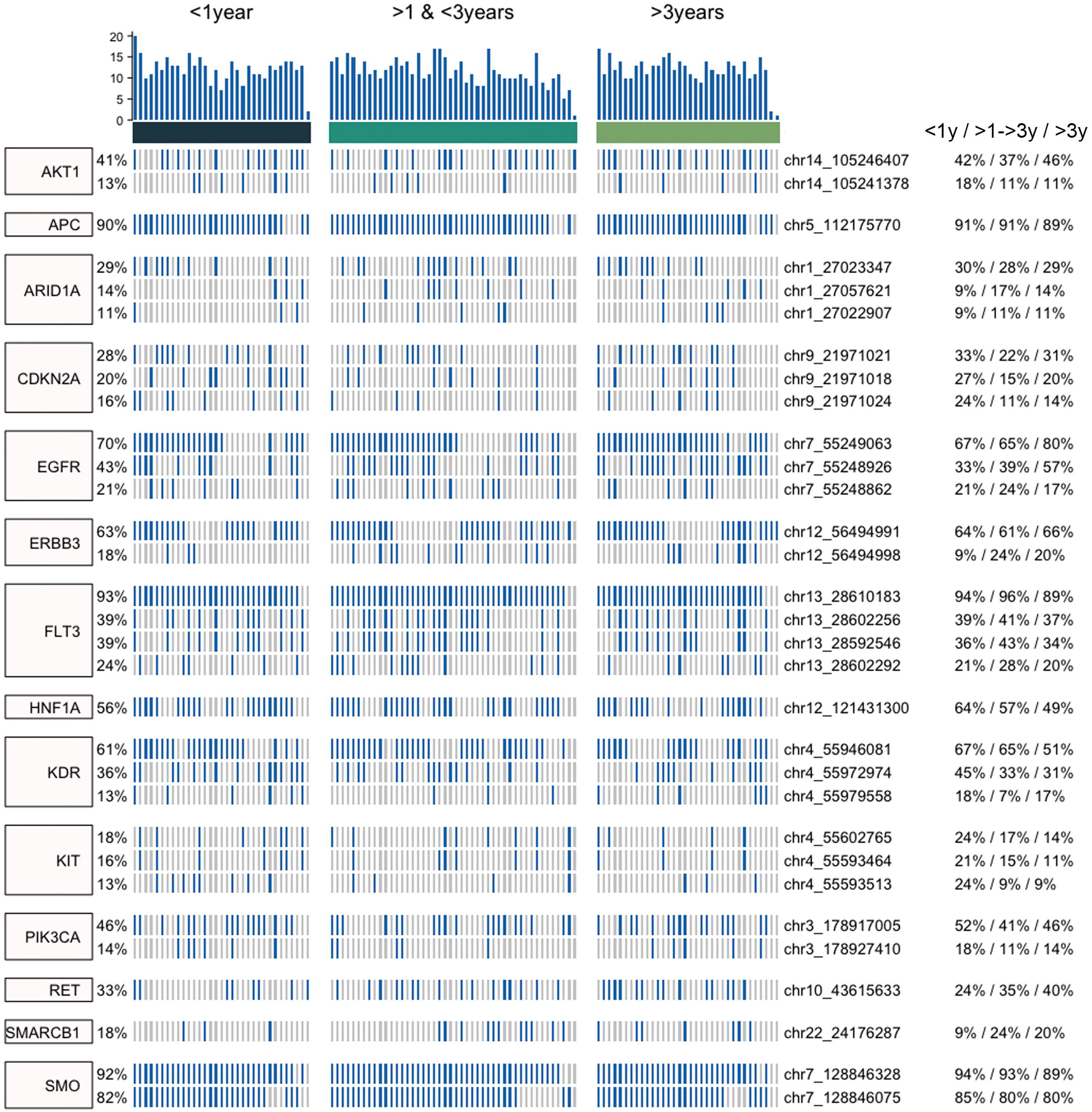

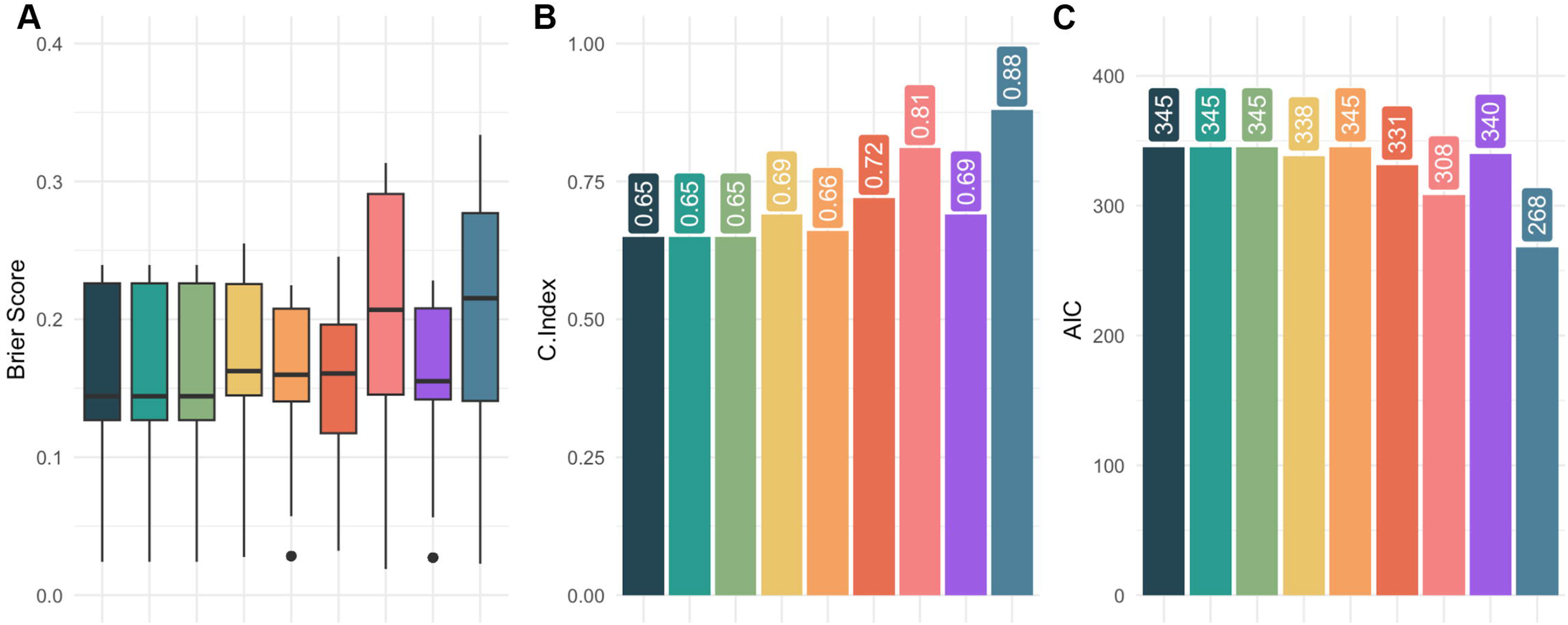

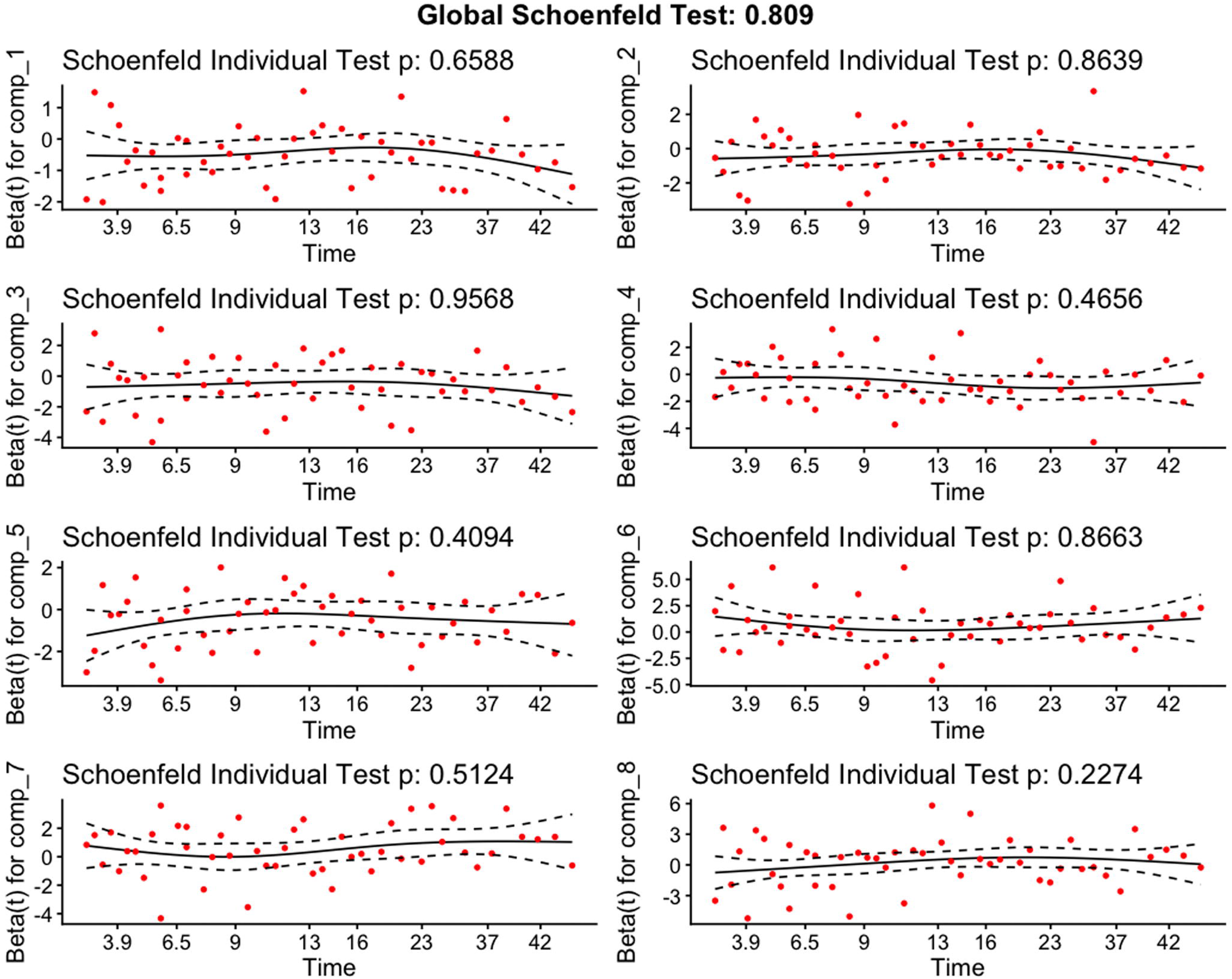

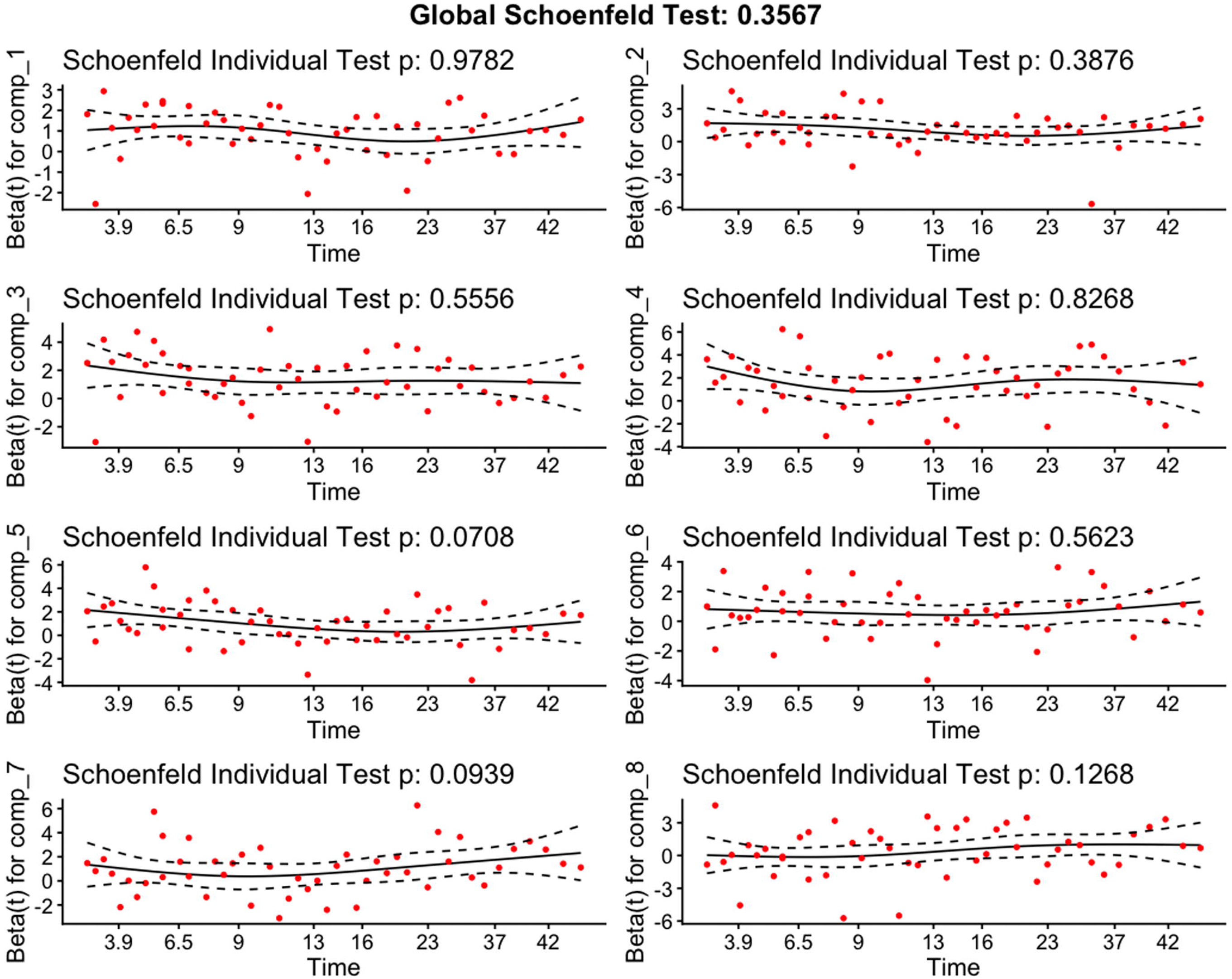

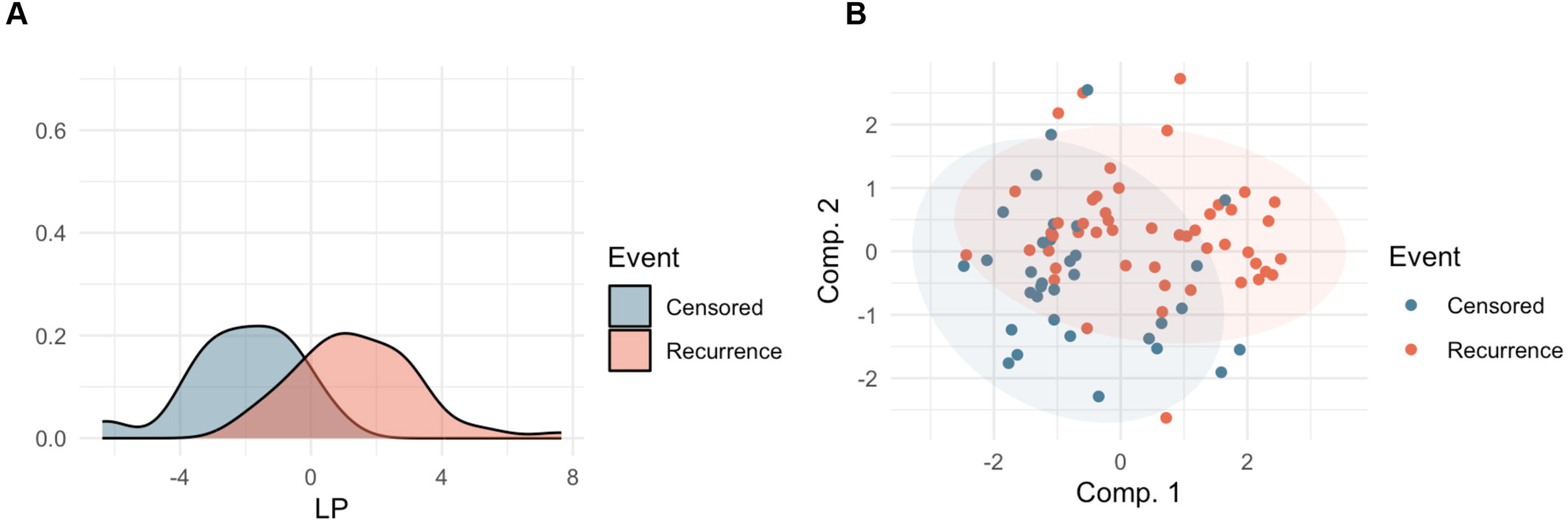

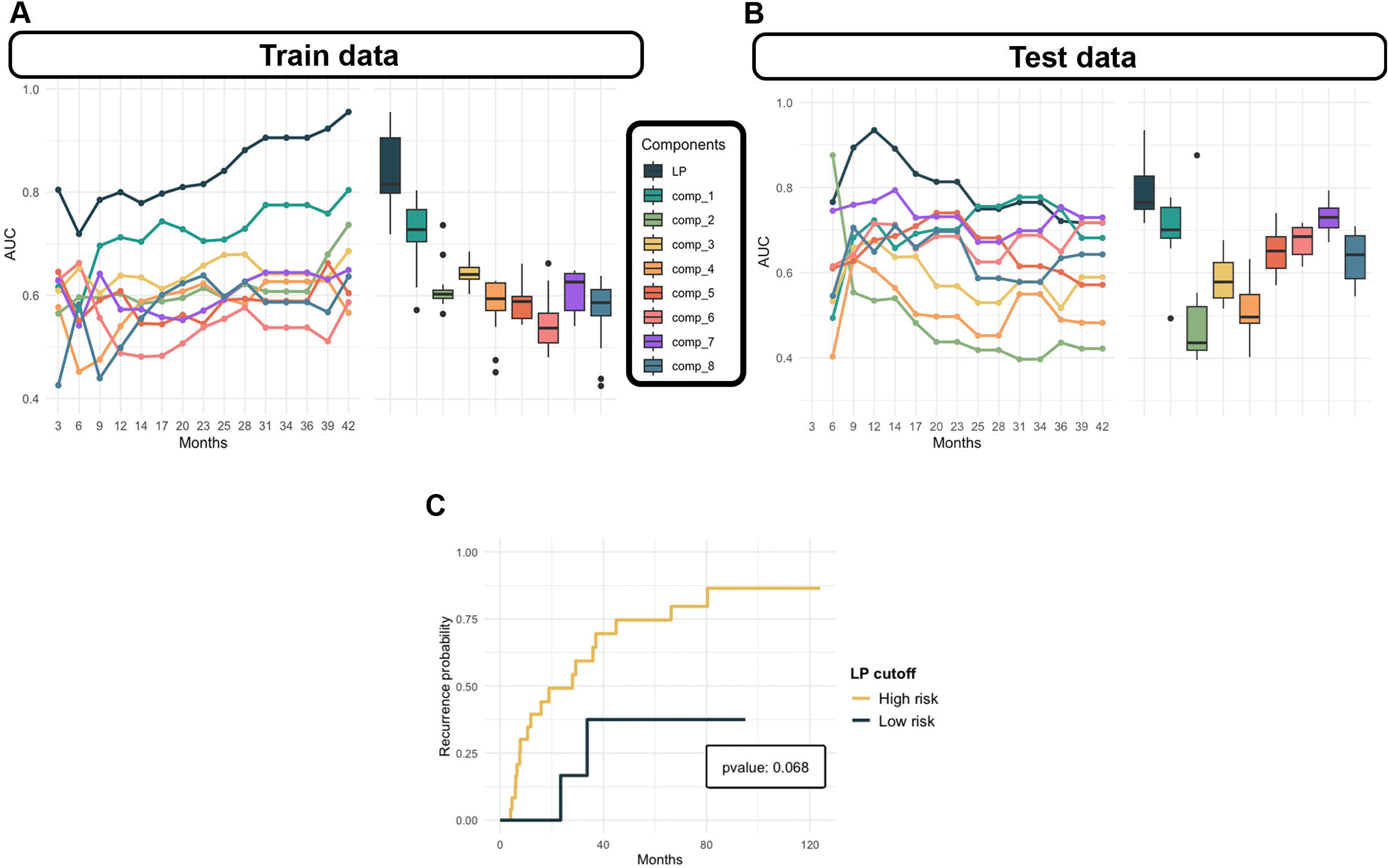

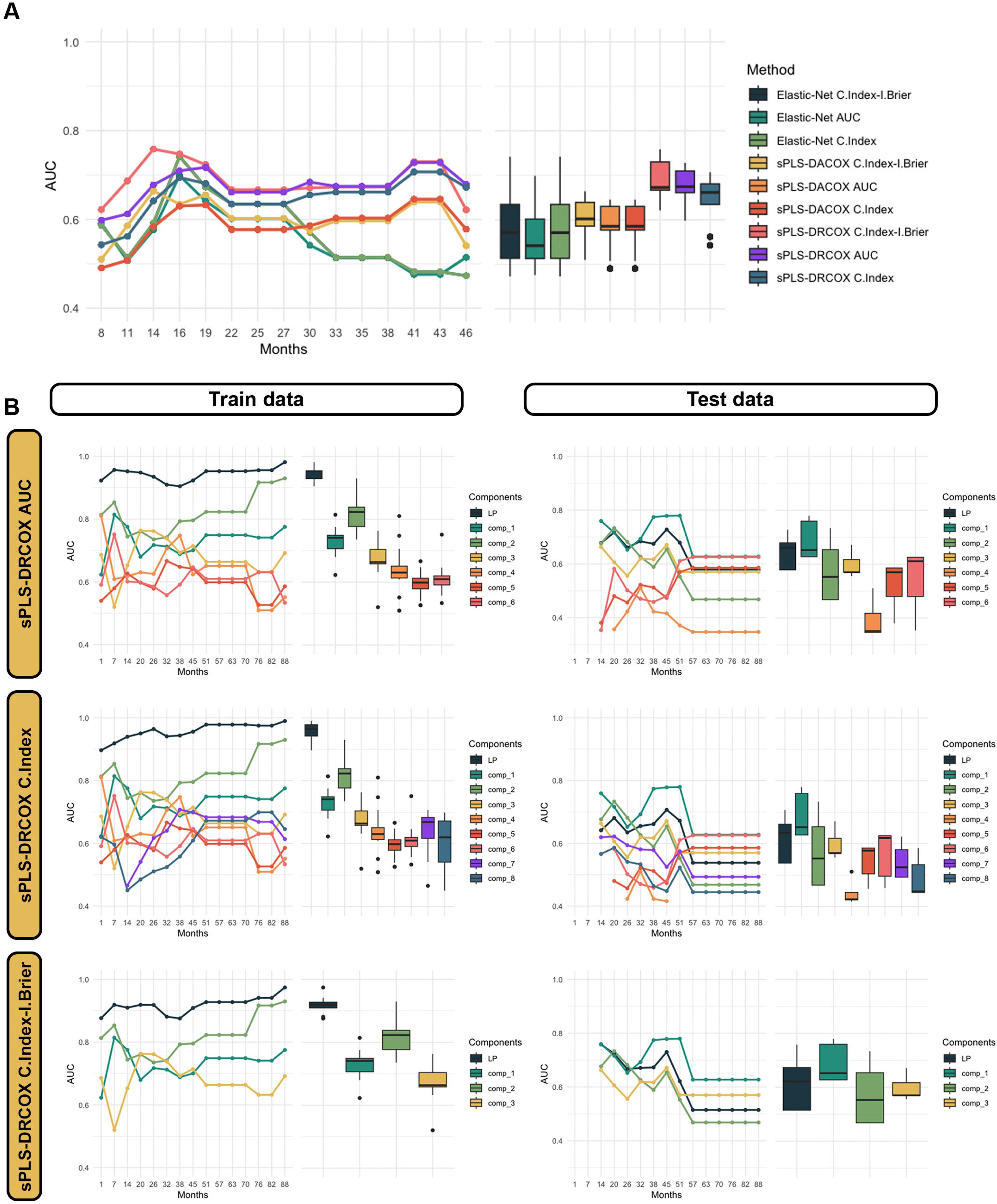

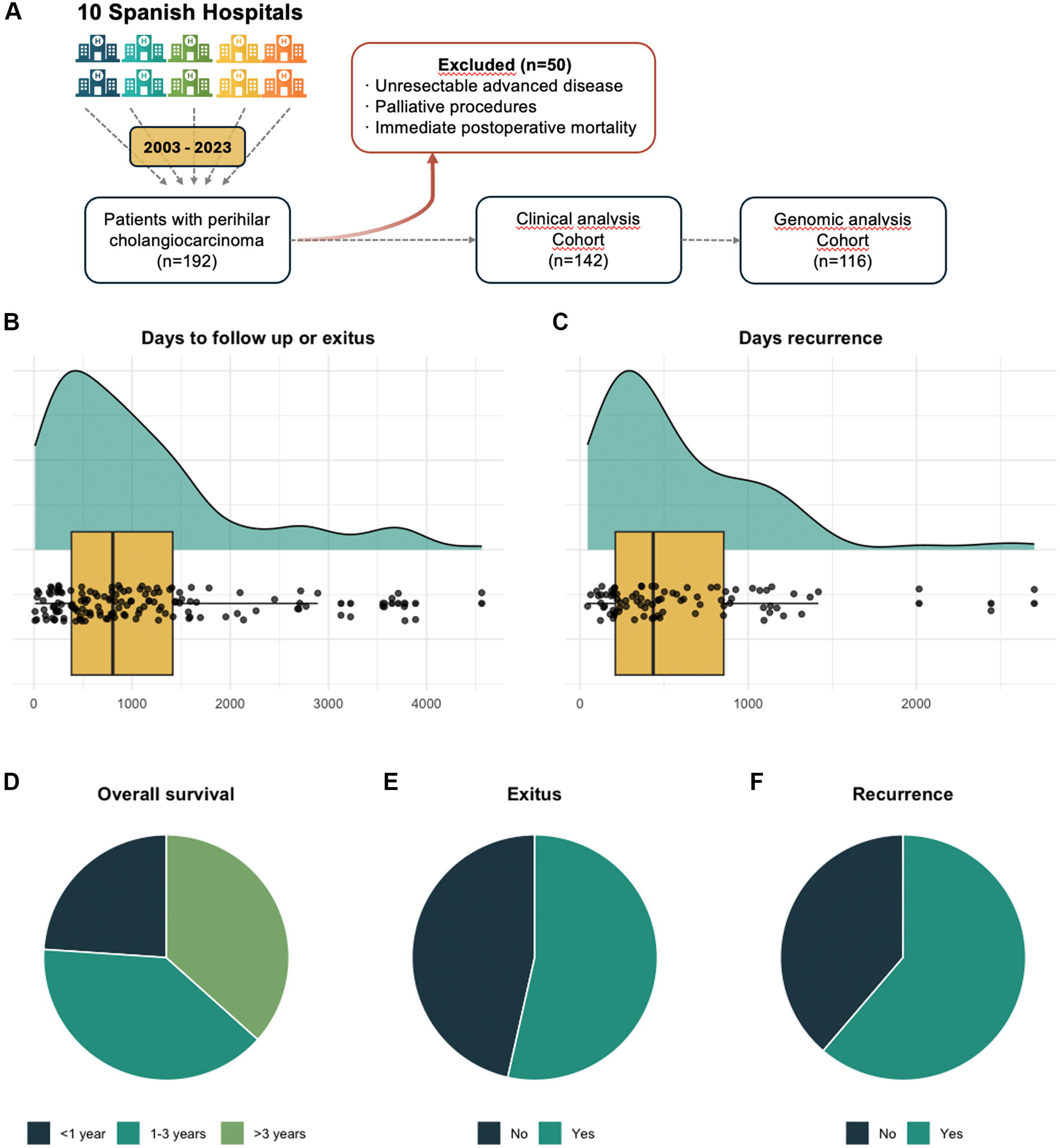

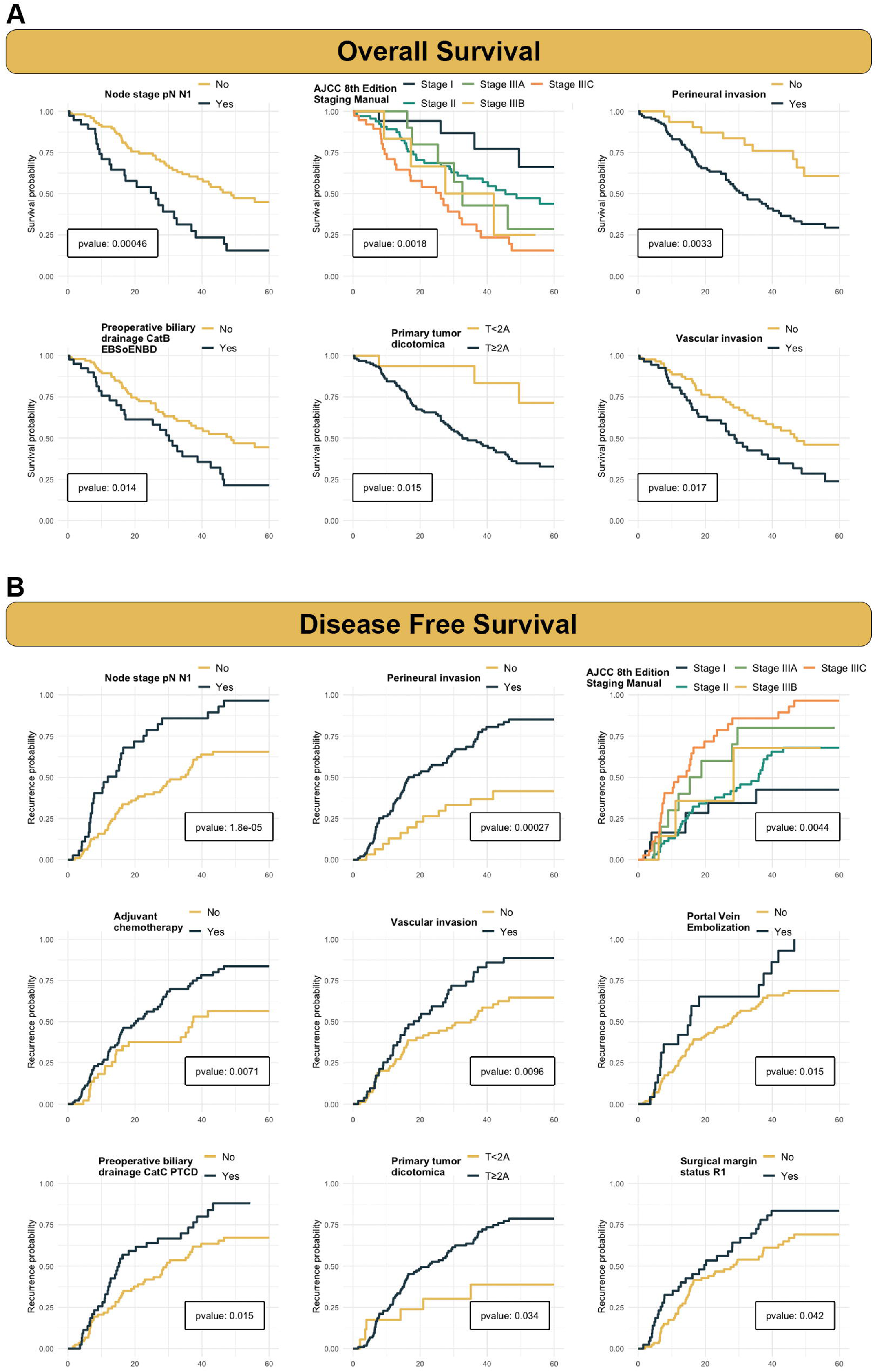

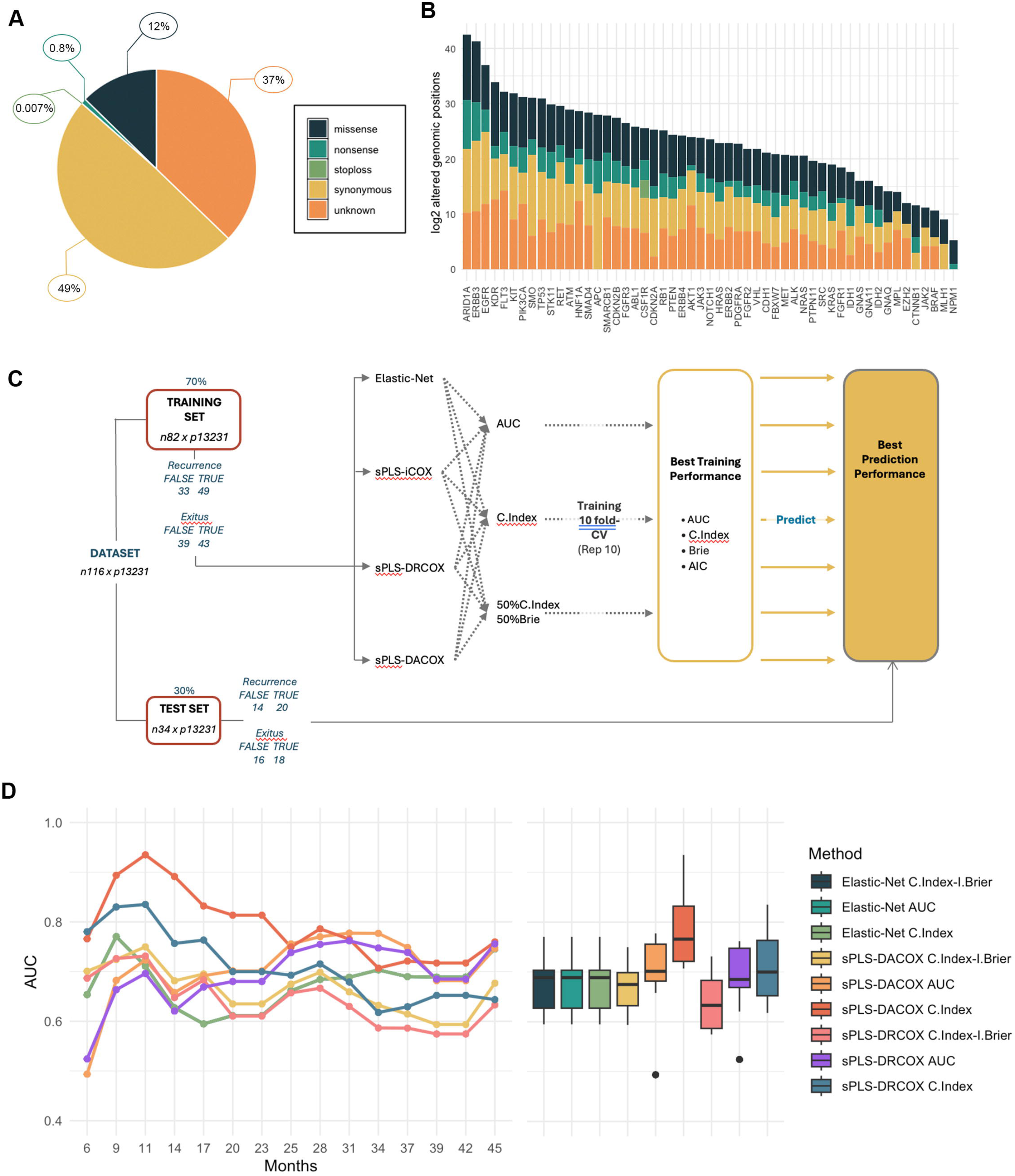

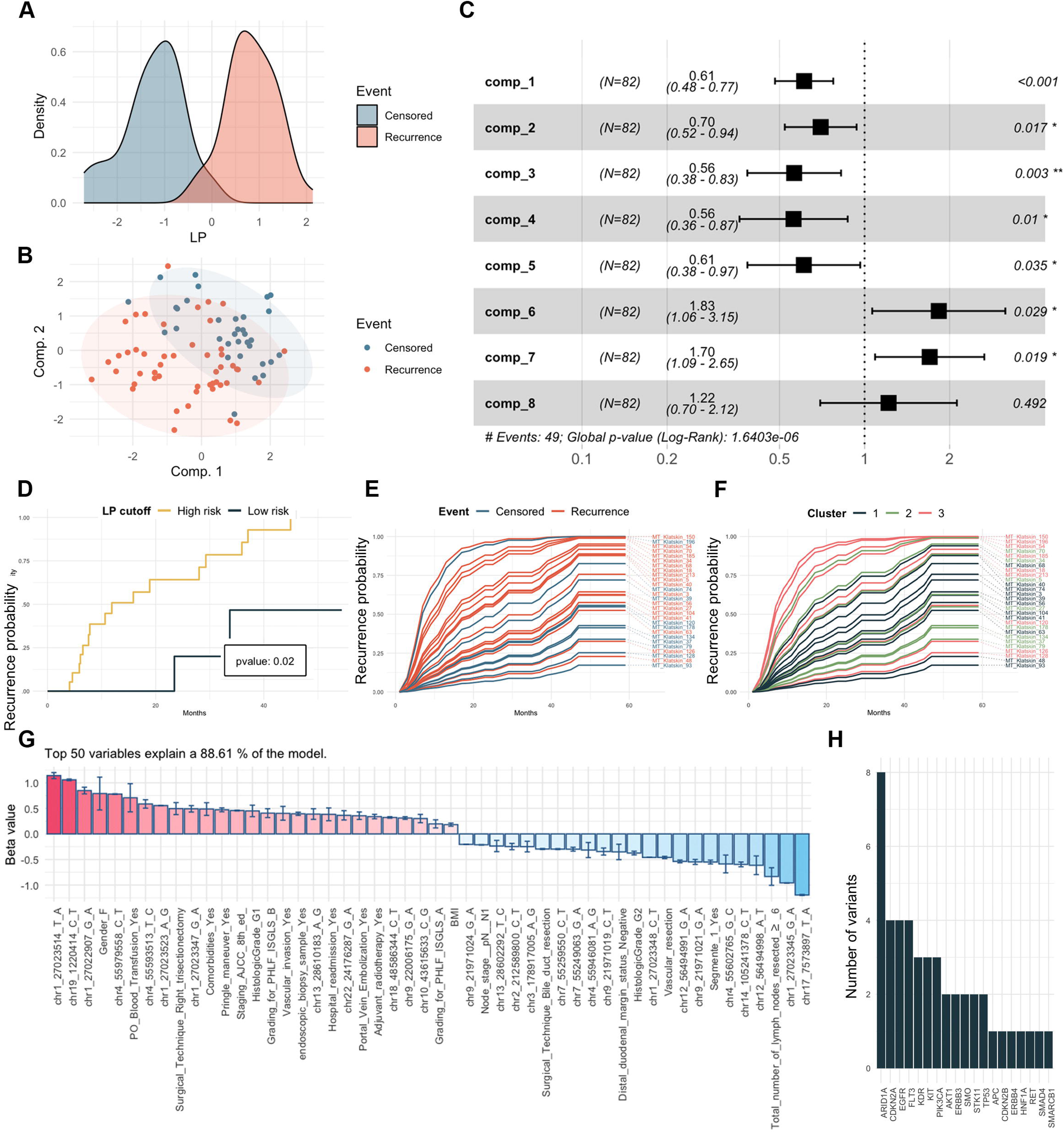

## Data Availability

Variant call data generated in this study have been deposited in the European Variation Archive (EVA) and are accessible under project accession PRJEB107911.
The rest of data, analytical methodologies, and research materials that support the findings of this study are available from the corresponding author upon reasonable request.

https://www.ebi.ac.uk/eva/?eva-study=PRJEB107911

## Author’s contributions

V.L-L., R.R., A.B-M. and F.L-R. contributed to the conception and design of the study, participated in data interpretation, and critically reviewed the manuscript. A.B-M., V.L-L., D.V-C., AI.A-G., CM.M-M. and F.L-R. performed the experimental work and statistical analyses. F.L-R. developed and conducted the bioinformatics analyses. All remaining authors contributed to patient inclusion and to the collection, processing and validation of clinical samples at their respective institutions. All authors read and approved the final version of the manuscript and take responsibility for the integrity of the data.

## Data availability statement

Variant call data generated in this study have been deposited in the European Variation Archive (EVA) [1] and are accessible under project accession PRJEB107911.

The rest of data, analytical methodologies, and research materials that support the findings of this study are available from the corresponding author upon reasonable request.

## Declaration of generative AI and AI-assisted technologies in the writing process

During the preparation of this work the author(s) used ChatGPT in order to improve grammar, spelling and the appearance of the text. After using this tool/service, the author(s) reviewed and edited the content as needed and take(s) full responsibility for the content of the publication.

## Abbreviations

AJCC: American Joint Committee on Cancer
AKT1: AKT serine/threonine kinase 1
APC: APC regulator of Wnt signaling pathway
ARID1A: AT-rich interaction domain 1A
AUC: area under the curve
CA 19-9: carbohydrate antigen 19-9
CDKN: cyclin dependent kinase inhibitor
COXMOS: Cox MultiBlock Survival
DIANA: DIvisive ANAlysis
DFS: Disease-free survival
EGFR: epidermal growth factor receptor
ERBB: Erb-B2 receptor tyrosine kinase
FLT3: Fms related receptor tyrosine kinase 3
IQR: interquartile range
KIT: KIT proto-oncogene, receptor tyrosine kinase
KRAS: KRAS proto-oncogene, GTPase
LP: linear predictor
OS: Overall survival
pCCA: Perihilar cholangiocarcinoma
PIK3: phosphatidylinositol-4,5-bisphosphate 3-kinase
PTCD: percutaneous transhepatic biliary drainage
PVE: portal vein embolization
RET: ret proto-oncogene
RTK: activated receptor tyrosine kinase
SMARCB1: SWI/SNF related BAF chromatin remodeling complex subunit B1
sPLS-DACOX: sparse Partial Least Squares Discriminant Analysis Cox
sPLS-DRCOX: sparse Partial Least Squares Deviance Residuals Cox
TP53: tumor protein P53
t-SNE: T-distributed Stochastic Neighbour Embedding

## Acknowledgements

We are particularly grateful for the collaboration of Biobank Network of the Region of Murcia, BIOBANC-MUR, registered on the Registro Nacional de Biobancos with registration number B.0000859. BIOBANC-MUR is supported by the “Instituto de Salud Carlos III (proyecto PT20/00109), by “Biomedical Research Institute of Murcia, (IMIB-Pascual Parrilla)” and by “Consejería de Salud de la Comunidad Autónoma de la Región de Murcia”. Likewise, authors would like to thank Genomics Core and Pathology Core of the IMIB for their technical assistance. We are sincerely grateful to Dr. Cuenca-Bermejo for his generous scientific guidance and for her meticulous contribution to the design and refinement of the figures, which greatly enhanced the clarity and presentation of our work.

