## supplementary data for "Integration of clinical and genomic data defines prognostic phenotypes in resected perihilar cholangiocarcinoma: a national multicenter study"

#### Supplementary Data Index


1. **Supplementary Methods**
   1.1. *Supplementary Methods 1. Study cohort and follow-up definitions*
   1.2. *Supplementary Methods 2. Univariate survival analyses*
   1.3. *Supplementary Methods 3. Unsupervised clustering methodology*
   1.4. *Supplementary Methods 4. Comparative statistical analyses of clinical variables*
   1.5. *Supplementary Methods 5. Tumour sampling and DNA extraction*
   1.6. *Supplementary Methods 6. Targeted sequencing panel and bioinformatic processing*
   1.7. *Supplementary Methods 7. Integrative clinical–genomic survival modelling (COXMOS framework)*
   1.8. *Supplementary Methods 8. Statistical software and computational environment*
2. **References**
3. **Supplementary Figures**
   3.1. *Supplementary Figure 1. Clinicopathological characteristics across DIANA-Gower clusters*
   3.2. *Supplementary Figure 2. Clinicopathological profile by DIANA-Gower cluster in the first 8 months*
   3.3. *Supplementary Figure 3. Variant-level alteration patterns stratified by recurrence status*

3.4. *Supplementary Figure 4. Variant-level alteration patterns stratified by overall survival groups*

3.5. *Supplementary Figure 5. Model selection metrics across candidate approaches*

3.6. *Supplementary Figure 6. Proportional hazards assumption assessment for the sPLS-DACOX Cox model*

3.7. *Supplementary Figure 7. Proportional hazards assumption assessment for the sPLS-DRCOX Cox model*

3.8. *Supplementary Figure 8. sPLS-DACOX recurrence model sample representation*

3.9. *Supplementary Figure 9. Time-dependent discrimination of latent components and full-model linear predictor*

3.10. *Supplementary Figure 10. OS model performance and component-level discrimination*

**4. Supplementary Tables**

4.1. *Supplementary Table 1*

4.2. *Supplementary Table 2*

4.3. *Supplementary Table 3*

4.4. *Supplementary Table 4*

4.5. *Supplementary Table 5*

4.6. *Supplementary Table 6*

4.7. *Supplementary Table 7*

### 1. Supplementary Methods

#### 1.1. Supplementary Methods 1. Study cohort and follow-up definitions

A multicentre, retrospective study was conducted including patients with perihilar cholangiocarcinoma (pCCA) who underwent surgical treatment between February 2003 and February 2023 across 10 Spanish centres: University Clinical Hospital Virgen de la Arrixaca (Murcia), University Hospital of Bellvitge and University Hospital of Vall d'Hebron (Barcelona), La Fe University Hospital and Clinical University Hospital (Valencia), General University Hospital Gregorio Marañón (Madrid), University Hospital Río Hortega (Valladolid), University Hospital Virgen del Rocío (Seville), Miguel Servet University Hospital (Zaragoza) and University Hospital of Badajoz (Badajoz). Patients without available recurrence data, those with unresectable advanced disease (AJCC 8th edition stages IVA-IVB), individuals treated with palliative procedures, those who experienced immediate postoperative mortality (Clavien-Dindo grade 5), and patients in whom an adequate tumour sample for genomic sequencing could not be obtained were excluded from the study. The complete study protocol, encompassing all predefined analytical methods and study procedures, was reviewed and approved by the Ethics Committee of Hospital Universitario Virgen de la Arrixaca (Murcia, Spain; registry number 2021-4-7-HCUVA), which acted as the central ethics committee, and was subsequently approved by the institutional review boards of all participating centres.

Baseline clinical characteristics, preoperative laboratory parameters (including bilirubin and CA 19-9), surgical details, and pathological findings (tumour differentiation grade, lymphovascular and perineural invasion, resection margin status, among others) were systematically recorded.

Oncological follow-up was performed from the date of surgery to the date of last contact or death. Overall survival (OS) was defined as the time from surgery to death from any cause or until the last date on which the patient was confirmed alive. Disease-free survival (DFS) was defined as the time from surgery to the first documented tumour recurrence, with patients without recurrence censored at their last known follow-up date.

##### 1.2. Supplementary Methods 2. Univariate survival analysis

Successive data subsets were constructed containing follow-up time, event status, and the variable under study, excluding those with only a single response level.

For each subset, a univariate Cox proportional hazards model was specified by creating a survival object using the *Surv()* function and fitting the model with *coxph()*, both from the survival package [1, 2]. Model coefficients were transformed into hazard ratios with 95% confidence intervals using the *tidy()* function from the broom package, systematically recording the term, HR, and p-value associated with each variable.

Variables with p-values ≤ 0.05 in the univariate Cox models were selected for detailed visualisation using Kaplan-Meier curves. Based on the survival data, category-stratified curves were estimated with *survfit(Surv() ~ variable)* from the survival package. The initial representation was generated using *ggsurvplot()*

from the survminer package [3], incorporating a risk table and p-value, and was subsequently refined and standardised using functions from the ggplot2 package.

##### 1.3. Supplementary Methods 3. Unsupervised clustering analysis

An unsupervised clustering analysis was performed using clinical and pathological variables. The dataset was curated by removing purely identifying variables, dates, follow-up-derived variables, and outcome variables (vital status and recurrence). Preoperative variables and intraoperative or surgical variables recorded during the procedure were selected for analysis.

All numerical and ordinal variables were standardised using the *scale()* function from the base R package stats to ensure comparable contributions to dissimilarity measures.

A Gower dissimilarity matrix was computed using the *daisy()* function from the cluster package with *metric* = *"gower"*. Based on this dissimilarity matrix *(diss* = *TRUE)*, a divisive hierarchical clustering model was fitted using the *diana()* function from the cluster package [4]. The final three-cluster solution was obtained using the *cutree()* and *as.hclust()* functions from the base stats package.

Cluster quality was assessed using the silhouette coefficient, computed with the *silhouette()* function from the cluster package, and visualised using *plot_silhouette()* [4]. The hierarchical structure and group separation were visualised with a dendrogram generated using *fviz_dend()* from the factoextra package [5].

The prognostic relevance of the clusters was evaluated using Cox proportional hazards models applied to datasets containing follow-up time truncated at 60 months, event status, and cluster assignment. Survival objects were defined using the *Surv()* function and models were fitted with *coxph()* from the survival package [1, 2]. Pairwise comparisons between clusters were performed using Tukey contrasts via the *glht()* function in combination with *mcp()* from the multcomp package [6], yielding adjusted p-values for differences in hazard. Cluster-stratified Kaplan-Meier curves were estimated with *survfit()* and visualised with *ggsurvplot()*.

##### 1.4. Supplementary Methods 4. Comparative analysis of clinical and pathological variables

Comparative analyses of clinical, analytical, surgical, and pathological variables were conducted according to: (i) OS categorised into three groups (<1 year, 1-3 years, and >3 years), (ii) dichotomous recurrence status (yes/no), and (iii) the three clusters obtained from the unsupervised clustering.

For trichotomised OS and for cluster comparisons, ordinal variables with ≥3 levels were analyzed using the Kruskal-Wallis test, followed by Dunn post hoc test with false discovery rate (FDR) correction when appropiate. Dichotomous categorical variables were analyzed using contingency tables and chi-square tests, applying Monte Carlo permutation tests (B = 100,000) when expected cell counts were <5. Pairwise comparisons were adjusted using the Benjamini-Hochberg procedure. Continuous variables were analysed after assessing normality with the Shapiro-

Wilk test. Normally distributed variables were analysed using one-way ANOVA with Tukey HSD post hoc tests, whereas non-normally distributed variables were analysed using Kruskal-Wallis tests with Dunn post hoc comparisons.

For dichotomous recurrence status, continuous variables were analysed using Welch's t-test or the Mann-Whitney test as appropriate. Dichotomous categorical variables were analysed using chi-square or Fisher's exact tests depending on expected cell counts.

##### 1.5. Supplementary Methods 5. Sample collection and DNA extraction

Formalin-fixed paraffin-embedded (FFPE) tumour tissue blocks were retrieved from pathology archives. For each case, three consecutive 10-µm sections were cut under sterile conditions. DNA was extracted using the QIAamp DNA FFPE Tissue Kit (Qiagen) following the manufacturer's protocol.

##### 1.6. Supplementary Methods 6. Targeted sequencing and bioinformatic analysis

Genomic profiling was performed using tumour DNA extracted from FFPE samples. Quantification of DNA and RNA was performed using Qubit® dsDNA (Thermofisher Scientific). Library preparation, sequencing and primary data analysis were performed by the Genomics Core of the Biomedical Research Institute of Murcia (IMIB).

The sequencing panel was designed using Ion AmpliSeq Designer v7.62, combining 207 cancer-associated hotspot regions included in the commercial Ion AmpliSeq™ Cancer Hotspot Panel v2 with the full coding exons (CDS) of ARID1A, ERBB3, and CDKN2B. The customised panel comprised 210 targets covering approximately 44.95 kb of the human genome reference hg19, generating 431 amplicons between 125 and 175 bp and achieving >99% coverage of target regions. Libraries were prepared through multiplex AmpliSeq™ PCR using the Ion AmpliSeq™ Library Kit 2.0 (Ion Torrent) according to manufacturer recommendations.

Template preparation was automated using the Ion 550 Kit-Chef (Ion Torrent), and sequencing of properly amplified Ion Sphere Particles (ISPs) was performed on the Ion GeneStudio™ S5 System (Ion Torrent) with Ion 550™ chips, yielding read numbers consistent with recommended specifications. Only samples meeting internal quality criteria were retained.

Primary bioinformatic processing was conducted with Torrent Suite v5.14.0 (Ion Torrent) independently for each sample. The workflow included: (i) removal of adapter and barcode sequences; (ii) alignment of reads to the human reference genome hg19 using the Torrent Mapping Alignment Program (TMAP); (iii) somatic variant calling using the Variant Caller module to generate unannotated VCF files; and (iv) functional and clinical annotation of variants with Ion Reporter v5.18.4.0, producing annotated VCF files subsequently used for downstream modelling.

Annotated per-sample VCF files were then processed and merged to generate a unified variant-by-sample matrix. Relevant fields were standardised, VCFs were compressed with bgzip and indexed with bcftools index. All sorted and indexed VCFs were merged using bcftools merge with a file list, enforcing inclusion of all samples (--force-samples), retaining all variants (-m all), and assigning reference genotypes to missing calls (--missing-to-ref). The merged VCF was subsequently indexed.

The merged VCF was converted into a tabular genotype matrix with one row per variant and one column per sample, containing genotype calls (0/0, 0/1, 1/1, etc.). This table was imported into R and filtered to retain only single-nucleotide variants by requiring REF and ALT alleles of length 1. Genotypes were recoded into a binary format: 0/0 (homozygous reference) was coded as 0, and any non-reference genotype (heterozygous or homozygous alternate) was coded as 1. A unique variant identifier was created by concatenating chromosome, position, reference allele, and alternate allele (CHROM_POS_REF_ALT). The resulting binary variant × sample matrix was used as the genomic input block for integrative survival models.

##### 1.7. Supplementary Methods 7. Integrative clinical-genomic survival models

To integrate genomic information with clinical variables in prognostic modelling, DFS was used as the primary endpoint. Predictors included (i) the binary variant-by-sample matrix derived from the merged VCF files and (ii) the selected clinical, analytical, surgical, and pathological covariates.

Somatic variants were used to generate an oncoprint summarising the distribution of alterations across patients. The alteration matrix (variant/locus × sample), together with locus-to-gene annotation, was formatted for oncoprint plotting and filtered to retain recurrent events. The oncoprint was created using the ComplexHeatmap package [7, 8], with columns annotated and split according to recurrence status or patient status and rows grouped by the corresponding gene when applicable.

Analyses were implemented in R using the COXMOS framework (Cox Multi-Omics Survival) [9], which enables penalised Cox modelling and latent-component approaches across multi-block datasets. The cohort with genomic information was randomly divided into two mutually exclusive subsets: a training set (70% of patients) and an independent test set (30%), using a fixed random seed (*set.seed(1234)*). All model development steps were performed exclusively in the training set; the test set was used solely for external evaluation.

Four families of Cox models were fitted within COXMOS: (1) elastic-net penalised Cox regression, combining L1 and L2 penalties; (2) sPLS-ICOX (Sparse Partial Least Squares for independent Cox), which constructs sparse latent components from the predictors; (3) sPLS-DACOX (sparse PLS double-additive Cox), which models additive contributions from clinical and genomic blocks; and (4) sPLS-DRCOX (sparse PLS dynamic relative-risk Cox), which allows time-varying effects of latent components. Up to eight latent components were allowed for PLS-based models, and penalisation and sparsity parameters were tuned for each method.

Hyperparameter optimisation was performed using 10-fold cross-validation repeated 10 times in the training set. Three optimisation criteria were evaluated for each model family: (i) maximisation of the time-dependent AUC over a predefined follow-up interval; (ii) maximisation of Harrell's concordance index

(C-index); and (iii) optimisation of a composite criterion averaging 50% C-index and 50% integrated Brier score, capturing both discrimination and calibration. A final model configuration was selected for each method-criterion combination.

Model performance was then assessed in the independent test set using the same metrics as in cross-validation, computed via the *eval_Coxmos_models()* function: time-dependent AUC, C-index, integrated Brier score, and Akaike information criterion (AIC), all derived from the model-specific linear predictors. For each model, the linear predictor (LP) was calculated for every patient and used as a univariate summary of recurrence risk.

To investigate the internal structure of component-based models, the contribution of each latent component to the risk signal was examined. Univariate Cox models were fitted with each component as the sole predictor, estimating hazard ratios of each component using *plot_forest()* and corresponding C-indices, while time-dependent AUCs were computed using *plot_evaluation()*. This allowed assessment of how prognostic information was distributed across components within the sPLS models.

The proportional hazards assumption was evaluated for all selected configurations using Schoenfeld residuals via the *cox.zph()* function within *plot_proportionalHazard()*. Both global and component-specific tests were examined, along with inspection for the absence of systematic temporal trends in residuals.

To derive clinically interpretable stratification rules, LP-based cut-points were obtained using the *getAutoKM()* function. These thresholds were estimated exclusively in the training set by identifying the LP values that maximised separation between survival curves (log-rank test). The resulting cut-points were applied without modification to the test set to generate high- and low-risk groups. Survival curves for these groups were estimated using *survfit()* from survival package [1, 2] and compared using the log-rank test.

To facilitate biological interpretation, regression coefficients (pseudo-betas) corresponding to original variables within the selected components were extracted using *plot_pseudobeta()*. These coefficients were used to compile descriptive lists of clinical and genomic variables with the highest absolute contributions to the latent risk components, providing insight into the underlying clinical-genomic signatures.

Additionally, analogous models were fitted using OS as the endpoint, following the same workflow (training/test split, cross-validation, and external evaluation). These models were used in an exploratory capacity and underwent the same methodological checks as the recurrence-oriented models.

##### 1.8. Supplementary Methods 8. Statistical environment

All analyses were performed in R version 4.4.0 (2024-04-24). Data manipulation was primarily conducted using dplyr and other packages within the tidyverse ecosystem [10], and model output was tidied and extracted using the broom package [11]. The fastDummies package [12] was used to create dummy variables, and the openxlsx package [13] was employed for importing and exporting Excel files. Figures and graphical outputs were generated using ggplot2 [14].

#### 3. Supplementary Figures

##### 3.1. Supplementary Figure S1. Clinicopathological characteristics across DIANA-Gower clusters.

Continuous variables are shown as boxplots with overlaid individual values (preoperative bilirubin and days to follow-up or death), and multi-level categorical variables are shown as proportional stacked bar charts (primary tumour category and AJCC 8th edition stage). Binary clinicopathological and surgical variables are displayed as proportional bar plots (Yes/No) across clusters, including comorbidities, bile duct resection, left/right hepatectomy, segmentectomy, Pringle manoeuvre, R1 margin status, vascular invasion, RBC transfusion, nodal status (pN1), histological grade (G1), lymphatic vessel invasion, perineural invasion, distal duodenal margin status (negative/positive with invasive cancer), and adjuvant chemotherapy. Statistical comparisons between clusters are indicated where applicable.

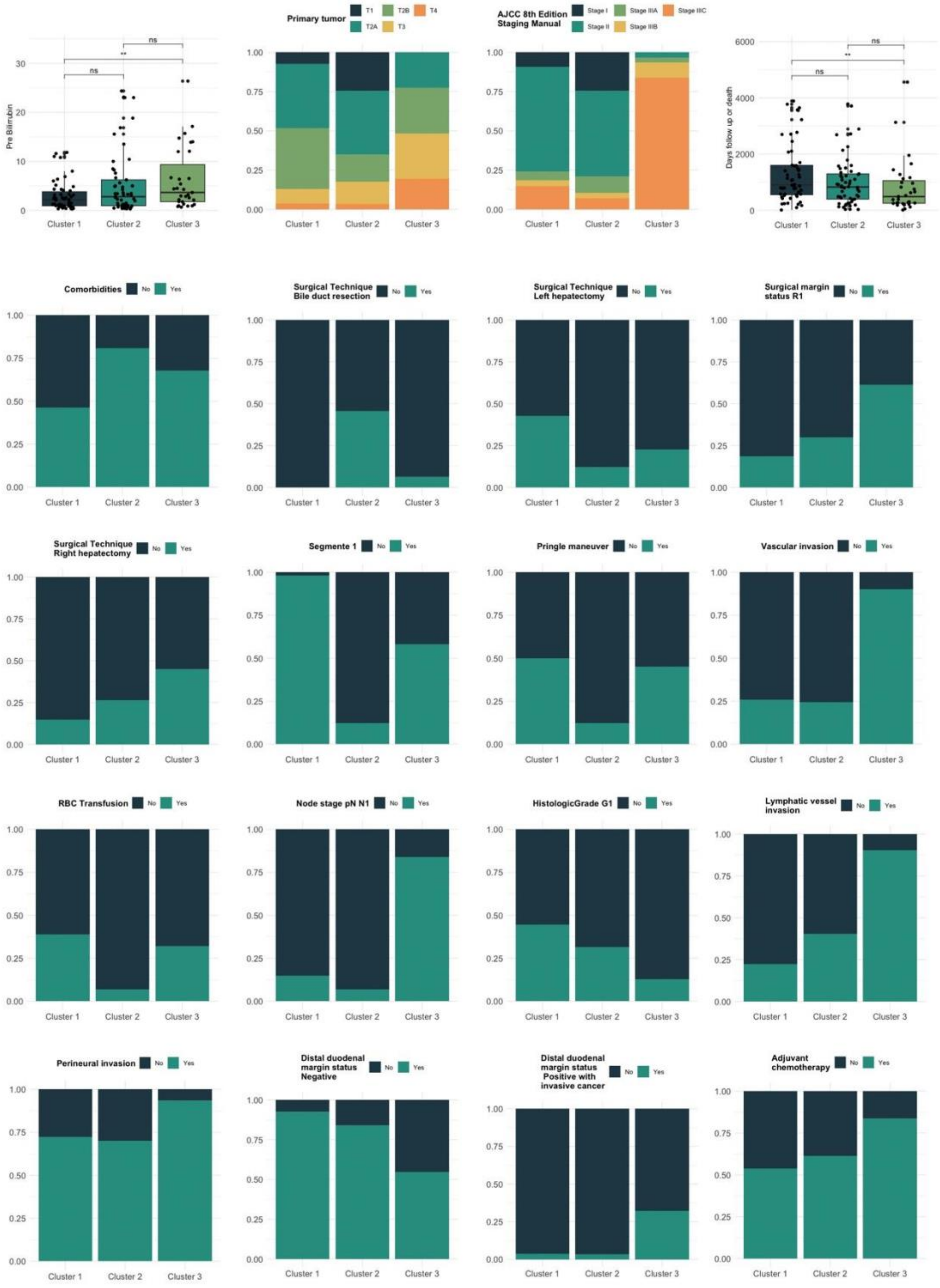
Pre Bilirrubin
ns
**
ns
Cluster 1
Cluster 2
Cluster 3
Primary tumor
T1
T2A
T2B
T3
T4
AJCC 8th Edition Staging Manual
Stage I
Stage II
Stage IIIA
Stage IIIB
Stage IIIC
Days follow up or death
Comorbidities
Surgical Technique Bile duct resection
Surgical Technique Left hepatectomy
Surgical margin status R1
Surgical Technique Right hepatectomy
Segmente 1
Pringle maneuver
Vascular invasion
RBC Transfusion
Node stage pN N1
HistologicGrade G1
Lymphatic vessel invasion
Perineural invasion
Distal duodenal margin status Negative
Distal duodenal margin status Positive with invasive cancer
Adjuvant chemotherapy
No
Yes

#### 3.2. Supplementary Figure S2. Clinicopathological profile by DIANA-Gower cluster in the first 8 months.

Proportional bar plots comparing clinicopathological and surgical variables across clusters among patients within 8 months, including AJCC 8th edition stage, ISGLS grade for post-hepatectomy liver failure (PHLF), bile duct resection, segmentectomy, nodal status (pN1), histological grade (G1), distal duodenal margin status (positive with invasive cancer), lymphatic vessel invasion, vascular invasion, and perioperative blood transfusion.

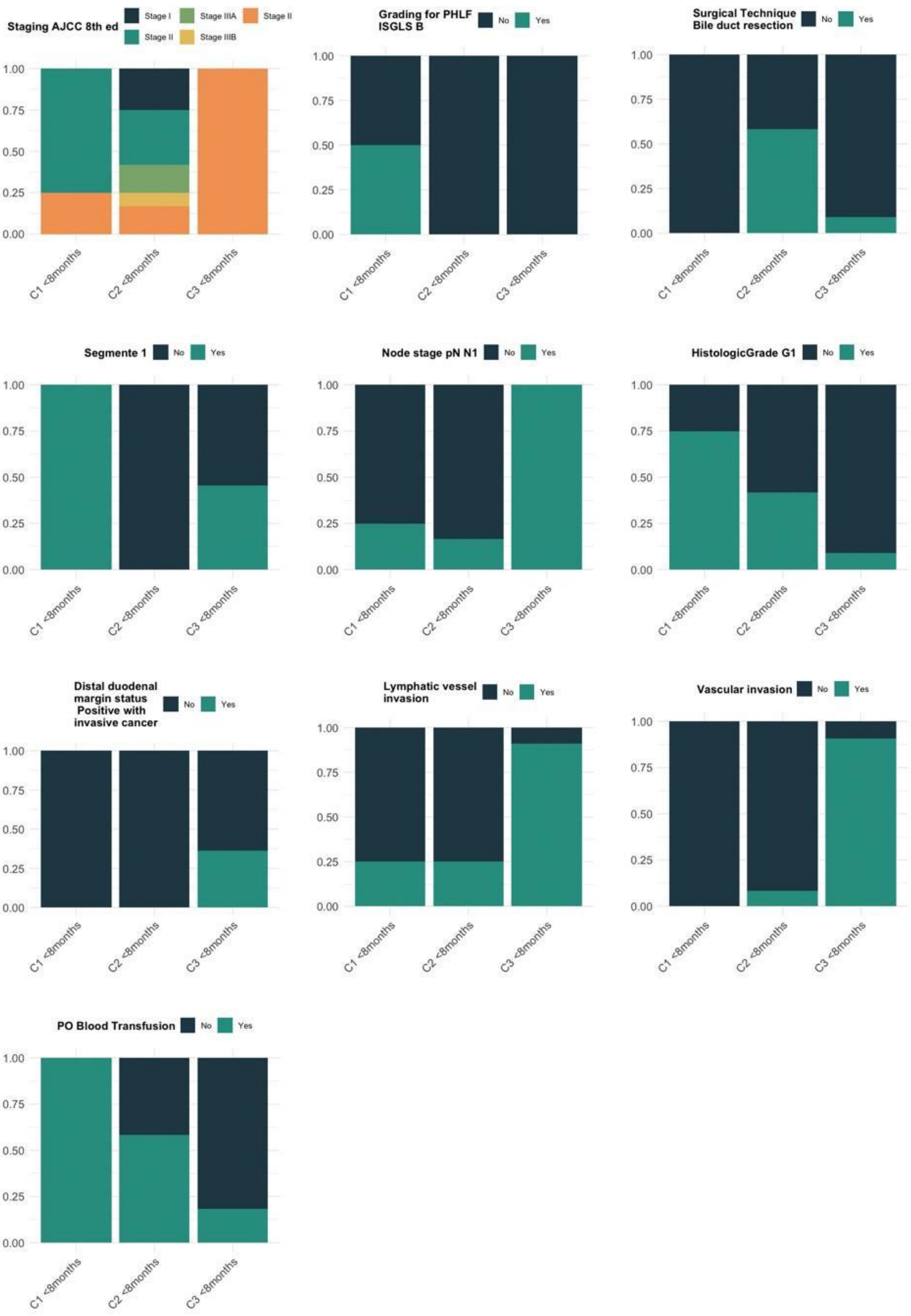

Staging AJCC 8th ed
Stage I
Stage IIIA
Stage II
Stage II
Stage IIIB
Grading for PHLF ISGLS B
Surgical Technique Bile duct resection
Segmente 1
Node stage pN N1
HistologicGrade G1
Distal duodenal margin status Positive with invasive cancer
Lymphatic vessel invasion
Vascular invasion
PO Blood Transfusion
No
Yes
0.00
0.25
0.50
0.75
1.00
C1 <8months
C2 <8months
C3 <8months

##### 3.3. Supplementary Figure S3. Variant-level alteration patterns stratified by recurrence status.

Oncoprint heatmap showing the presence/absence of selected variants grouped by gene across patients without recurrence versus with recurrence. The upper bar plots summarise the number of altered positions per patient. For each variant, the corresponding genomic coordinate is indicated, and the frequency of alteration is reported as non-recurrence / recurrence (%).

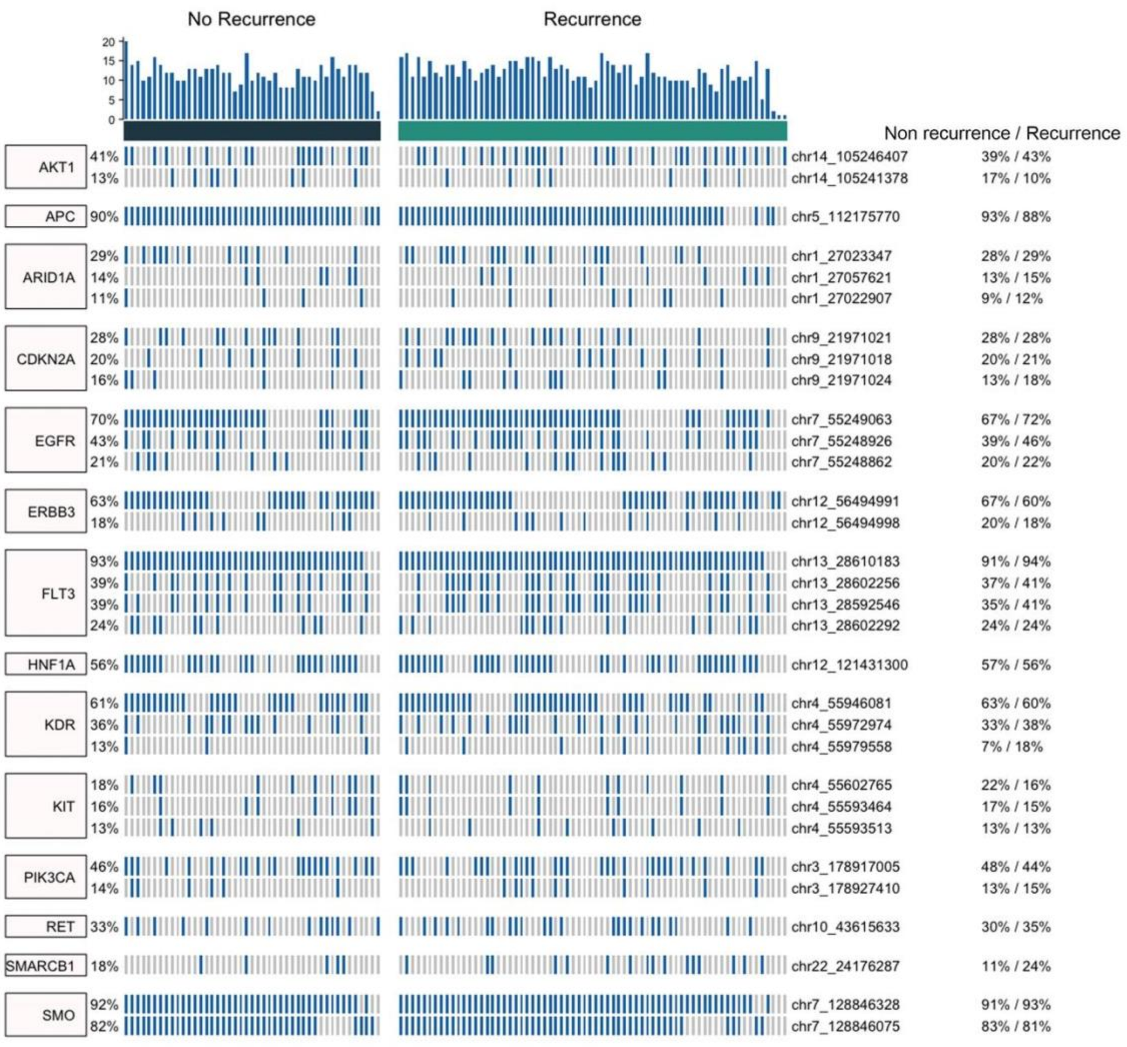

##### 3.4. Supplementary Figure S4. Variant-level alteration patterns stratified by overall survival groups.

Oncoprint heatmap showing the presence/absence of selected variants grouped by gene across patients stratified by overall survival (<1 year, 1–3 years, and >3 years). The upper bar plots summarise the number of altered positions per patient. For each variant, the corresponding genomic coordinate is indicated, and alteration frequencies are reported for each survival group (<1y / 1–3y / >3y, %).

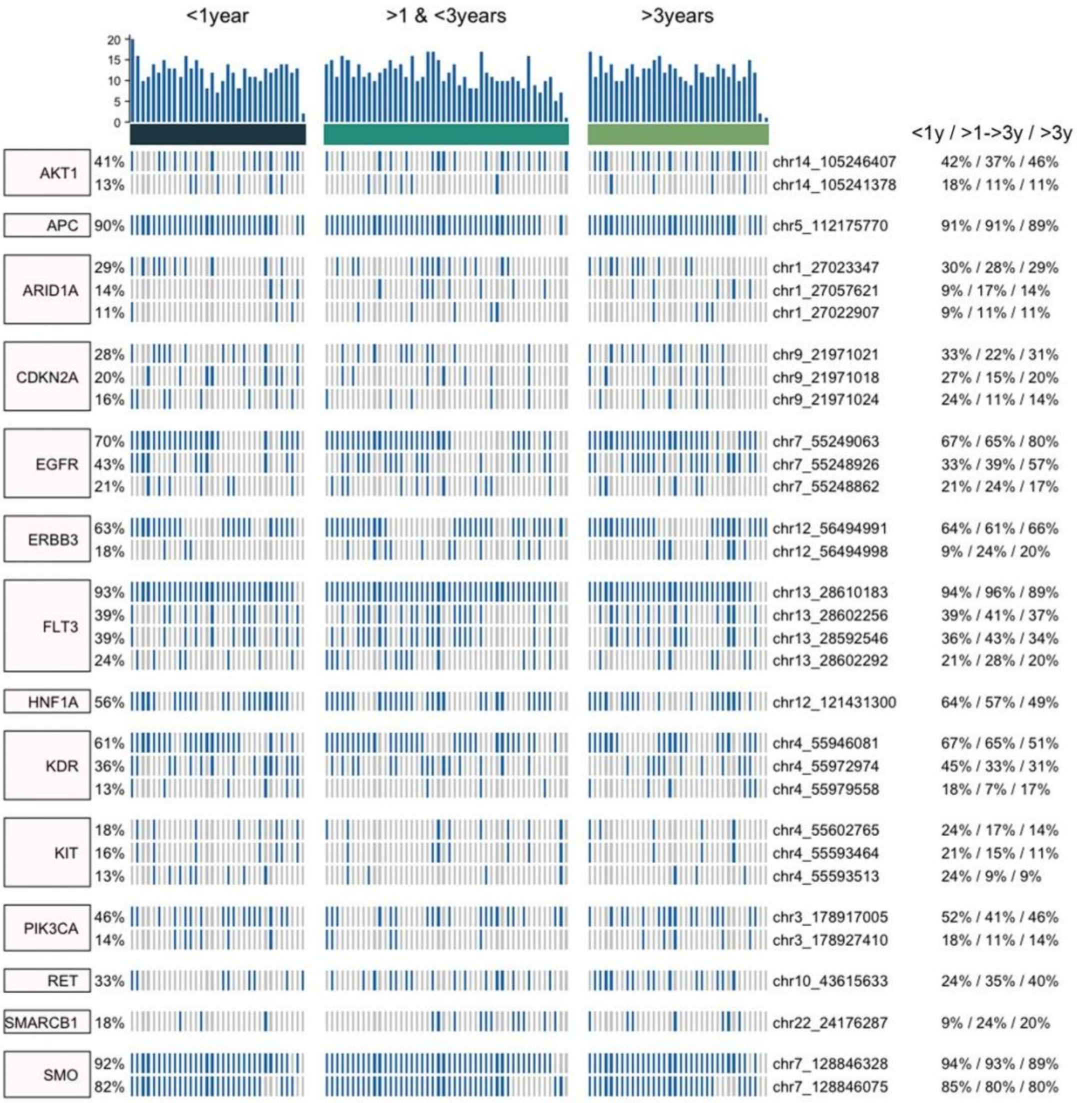
<1year
>1 & <3years
>3years
20
15
10
5
0
<1y / >1->3y / >3y
AKT1
41%
13%
chr14_105246407
chr14_105241378
42% / 37% / 46%
18% / 11% / 11%
APC
90%
chr5_112175770
91% / 91% / 89%
ARID1A
29%
14%
11%
chr1_27023347
chr1_27057621
chr1_27022907
30% / 28% / 29%
9% / 17% / 14%
9% / 11% / 11%
CDKN2A
28%
20%
16%
chr9_21971021
chr9_21971018
chr9_21971024
33% / 22% / 31%
27% / 15% / 20%
24% / 11% / 14%
EGFR
70%
43%
21%
chr7_55249063
chr7_55248926
chr7_55248862
67% / 65% / 80%
33% / 39% / 57%
21% / 24% / 17%
ERBB3
63%
18%
chr12_56494991
chr12_56494998
64% / 61% / 66%
9% / 24% / 20%
FLT3
93%
39%
39%
24%
chr13_28610183
chr13_28602256
chr13_28592546
chr13_28602292
94% / 96% / 89%
39% / 41% / 37%
36% / 43% / 34%
21% / 28% / 20%
HNF1A
56%
chr12_121431300
64% / 57% / 49%
KDR
61%
36%
13%
chr4_55946081
chr4_55972974
chr4_55979558
67% / 65% / 51%
45% / 33% / 31%
18% / 7% / 17%
KIT
18%
16%
13%
chr4_55602765
chr4_55593464
chr4_55593513
24% / 17% / 14%
21% / 15% / 11%
24% / 9% / 9%
PIK3CA
46%
14%
chr3_178917005
chr3_178927410
52% / 41% / 46%
18% / 11% / 14%
RET
33%
chr10_43615633
24% / 35% / 40%
SMARCB1
18%
chr22_24176287
9% / 24% / 20%
SMO
92%
82%
chr7_128846328
chr7_128846075
94% / 93% / 89%
85% / 80% / 80%

##### 3.5. Supplementary Figure S5. Model selection metrics across candidate approaches.

(A) Boxplots of the Brier score across candidate models. (B) Bar plot of concordance index (C-index) values for each model. (C) Bar plot of Akaike Information Criterion (AIC) values for each model.

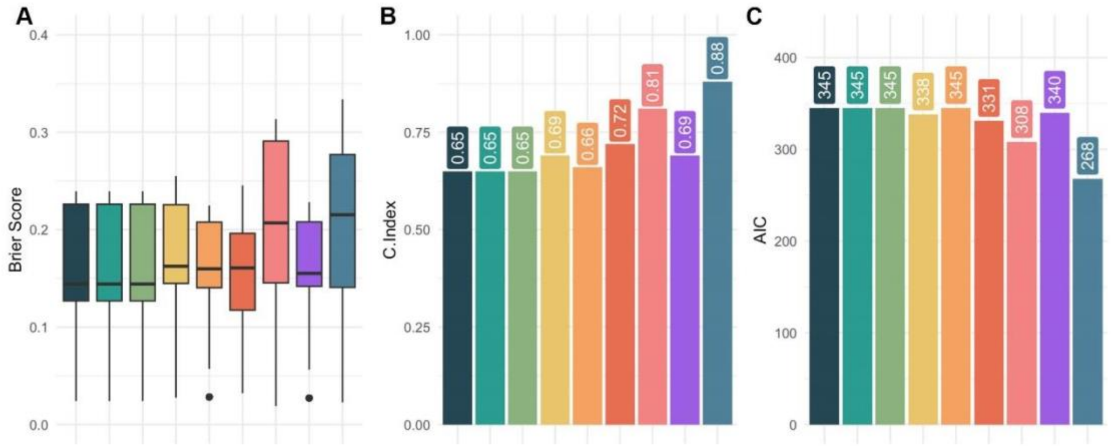

##### 3.6. Supplementary Figure S6. Proportional hazards assumption assessment for the sPLS-DACOX Cox model.

Schoenfeld residual plots for each latent component showing β(t) over time with fitted smooth curves and confidence bands, together with individual and global Schoenfeld test p-values.

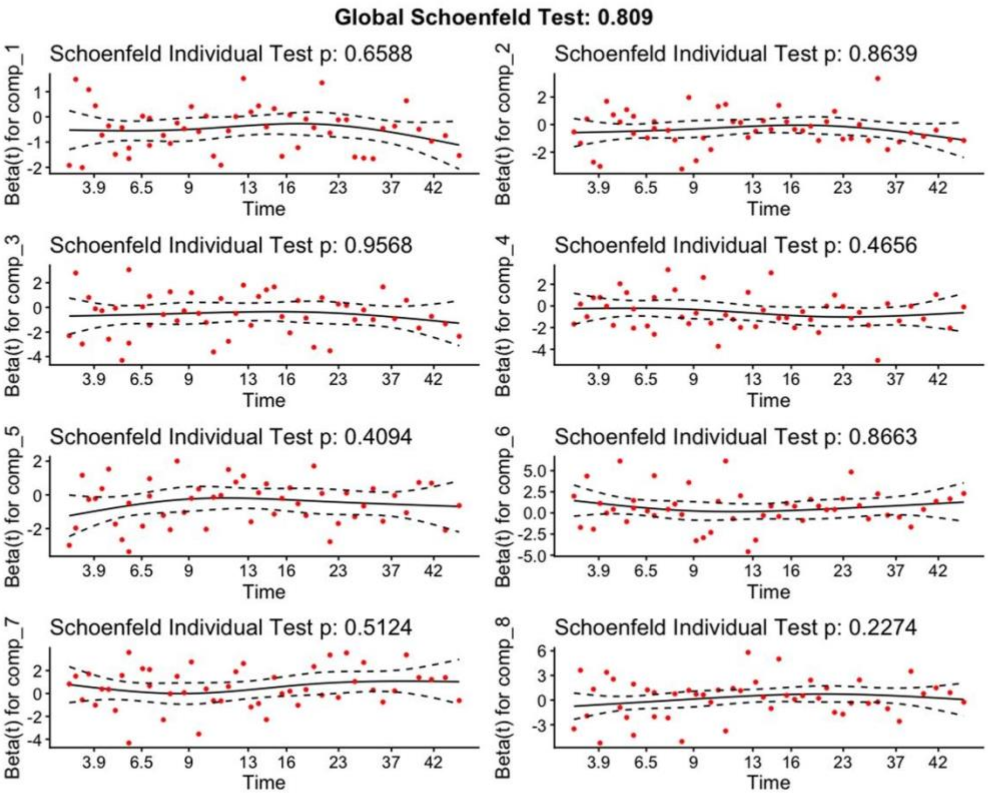

##### 3.7. Supplementary Figure S7. Proportional hazards assumption assessment for the sPLS-DRCOX Cox model.

Schoenfeld residual plots for each latent component showing β(t) over time with fitted smooth curves and confidence bands, together with individual and global Schoenfeld test p-values.

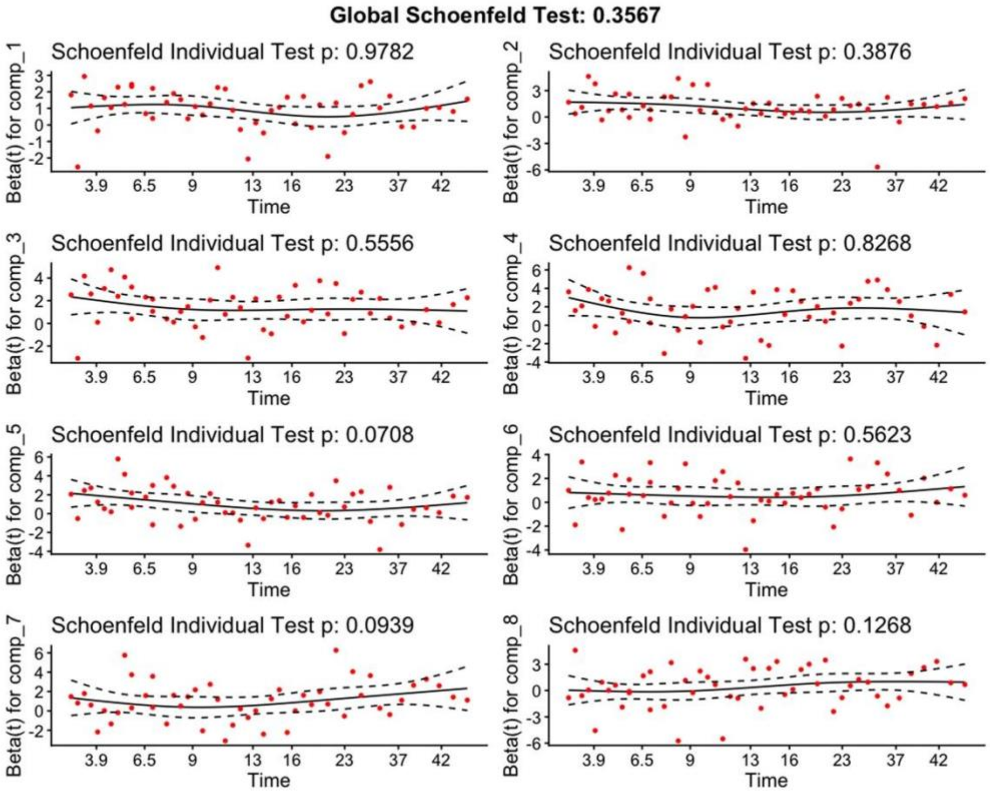

##### 3.8. Supplementary Figure S8. sPLS-DACOX recurrence model sample representation.

(A) Density distribution of the model-derived LP stratified by event status (censored vs recurrence). (B) Sample projection on the first two latent components, coloured by event status.

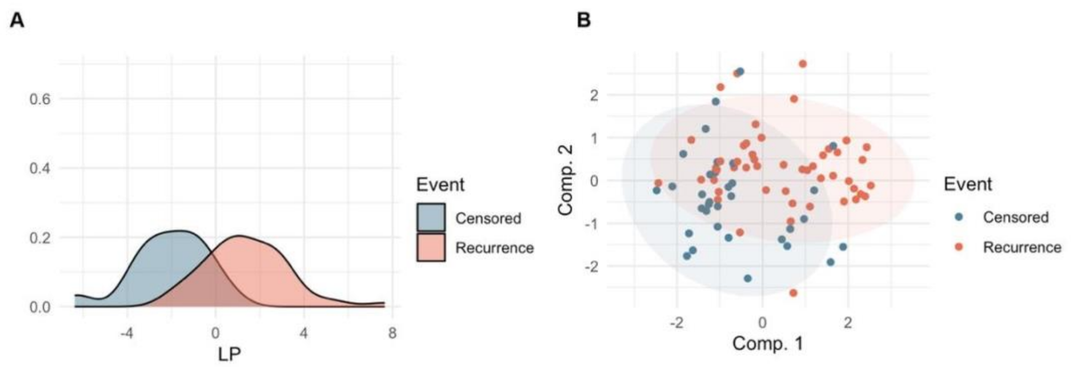

##### 3.9. Supplementary Figure S9. Time-dependent discrimination of latent components and full-model linear predictor.

(A) Training set: time-dependent AUC curves (months) for each latent component together with the LP from the full model, with boxplots summarising AUC distributions. (B) Test set: time-dependent AUC curves (months) for each latent component together with the full-model LP, with boxplots summarising AUC distributions. (C) Kaplan-Meier curve of recurrence probability in the test set stratified by full-model LP risk groups (high vs low risk), with log-rank p-value.

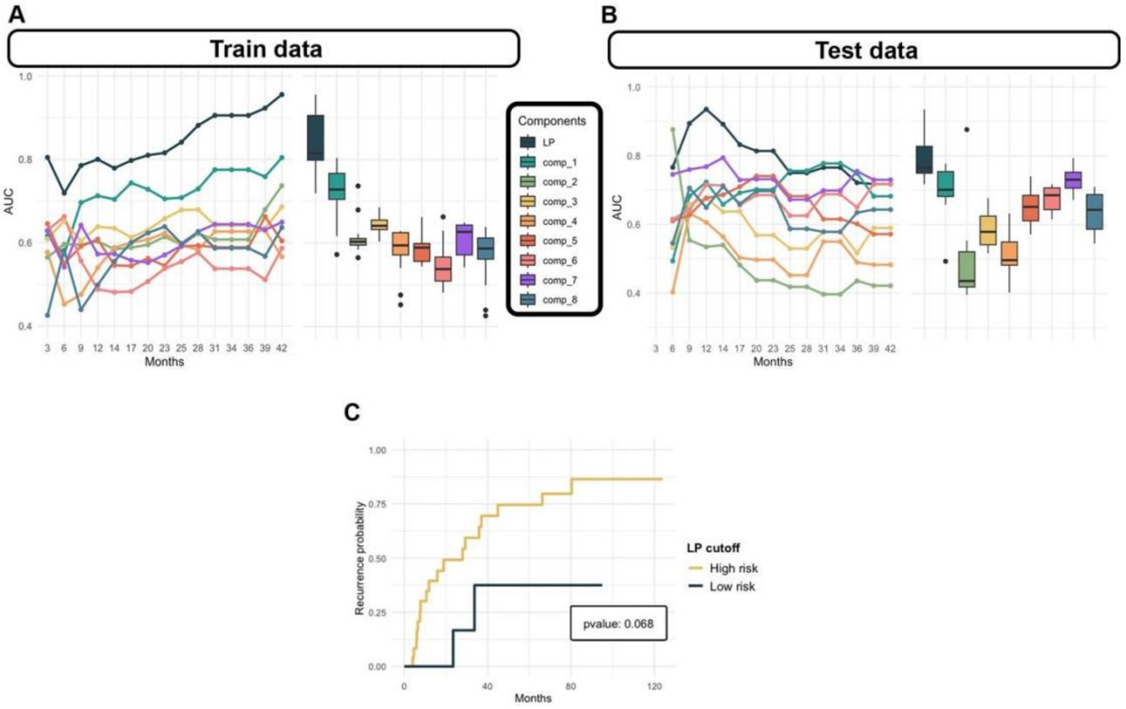

##### 3.10. Supplementary Figure S10. OS model performance and component-level discrimination.

(A) Time-dependent AUC profiles (months) and corresponding boxplots comparing OS prediction performance across Elastic-Net and sPLS-based models (sPLS-DACOX and sPLS-DRCOX) under different optimisation criteria. (B) For the OS sPLS-DRCOX models, time-dependent AUC curves (months) for the full-model LP together with the individual latent components, shown separately for the training and test sets, with boxplots summarising AUC distributions for each component and the LP.

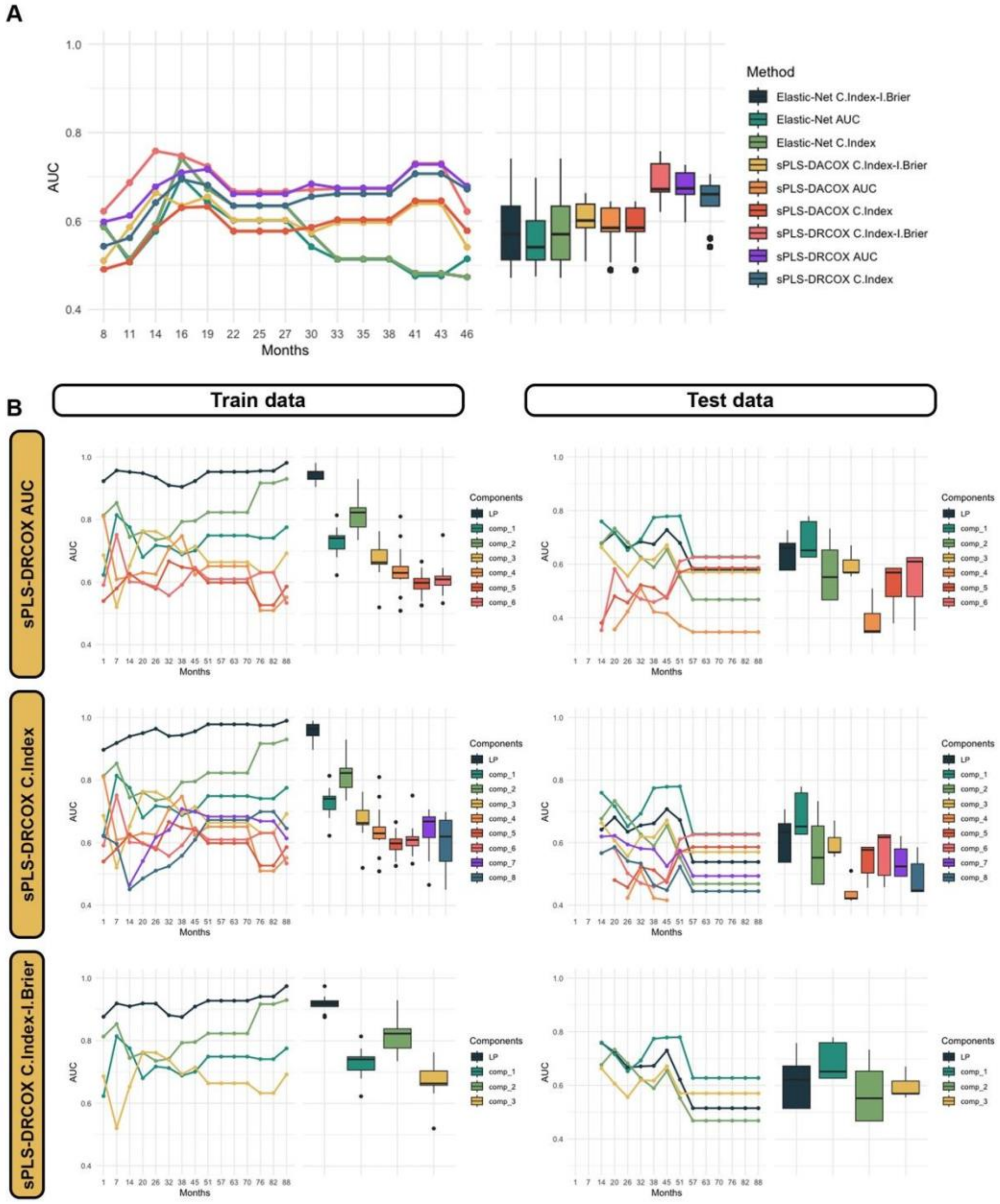
A
AUC
Months
Method
Elastic-Net C.Index-I.Brier
Elastic-Net AUC
Elastic-Net C.Index
sPLS-DACOX C.Index-I.Brier
sPLS-DACOX AUC
sPLS-DACOX C.Index
sPLS-DRCOX C.Index-I.Brier
sPLS-DRCOX AUC
sPLS-DRCOX C.Index
B
Train data
Test data
sPLS-DRCOX AUC
sPLS-DRCOX C.Index
sPLS-DRCOX C.Index-I.Brier
Components
LP
comp_1
comp_2
comp_3
comp_4
comp_5
comp_6
comp_7
comp_8

### 4. Supplementary Tables

#### *4.1. Supplementary Table 1*

| Variable | Test | p.value | post-hoc | post-hoc pvalue |
|---|---|---|---|---|
| **Age** | Kruskal-Wallis test | 0,58 | NS | NS |
| **BMI** | Kruskal-Wallis test | 0,52 | NS | NS |
| **Pre_Bilirrubin** | Kruskal-Wallis test | 0,93 | NS | NS |
| **Pre_Ca19_9** | Kruskal-Wallis test | 0,5 | NS | NS |
| **Primary_tumor** | Kruskal-Wallis Test | 0,81 | NS | NS |
| **Staging_AJCC_8th_ed_** | Kruskal-Wallis Test | 0,034 | Dunn test (FDR) | 1-3 years-<1 year (pvalue: 0.23) // >3 years-<1 year (pvalue: 0.032) // >3 years-1-3 years (pvalue: 0.17) |
| **ICU_days** | Kruskal-Wallis test | 0,99 | NS | NS |
| **Hospital_total_days** | Kruskal-Wallis test | 0,68 | NS | NS |
| **Vascular_resection** | Chi2 con permutación de Monte Carlo | 0,14 | NS | NS |
| **Primary_tumor_dichotomic** | Chi2 con permutación de Monte Carlo | 0,47 | NS | NS |
| **Staging_AJCC_8th_ed__dichotomic** | Chi2 test | 0,071 | NS | NS |
| **Clavien** | Kruskal-Wallis Test | 0,84 | NS | NS |
| **Clavien_dichotomic** | Chi2 test | 0,84 | NS | NS |
| **Gender_F** | Chi2 test | 0,89 | NS | NS |
| **Comorbidities_Yes** | Chi2 test | 0,026 | Benjamini-Hochberg (FDR) | 1_a_3_years-<_1_year (pvalue: 0.17) // >_3_years-<_1_year (pvalue: 0.039) // >_3_years-1_a_3_years (pvalue: 0.36) |
| **endoscopic_biopsy_sample_Yes** | Chi2 test | 0,36 | NS | NS |
| **Portal_Vein_Embolization_Yes** | Chi2 test | 0,23 | NS | NS |
| **Preoperative_biliary_drainage_CatB_EBSoENBD** | Chi2 test | 0,18 | NS | NS |
| **Preoperative_biliary_drainage_CatC_PTCD** | Chi2 test | 0,26 | NS | NS |
| **Surgical_Technique_Bile duct resection** | Chi2 test | 0,45 | NS | NS |
| **Surgical_Technique_Bile duct resection + 4b-5 segmentectomy** | Chi2 con permutación de Monte Carlo | 1 | NS | NS |
| **Surgical_Technique_Bile duct resection + vascular resection** | Chi2 con permutación de Monte Carlo | 0,056 | NS | NS |
| **Surgical_Technique_Left hepatectomy** | Chi2 test | 0,38 | NS | NS |

| | | | | |
|---|---|---|---|---|
| **Surgical_Technique_Left hepatectomy + vascular resection** | Chi2 con permutación de Monte Carlo | 0,12 | NS | NS |
| **Surgical_Technique_Left trisectionectomy** | Chi2 con permutación de Monte Carlo | 0,79 | NS | NS |
| **Surgical_Technique_Mesohepatectomy** | Chi2 con permutación de Monte Carlo | 1 | NS | NS |
| **Surgical_Technique_Right hepatectomy** | Chi2 test | 0,37 | NS | NS |
| **Surgical_Technique_Right hepatectomy + vascular resection** | Chi2 con permutación de Monte Carlo | 0,19 | NS | NS |
| **Surgical_Technique_Right trisectionectomy** | Chi2 con permutación de Monte Carlo | 0,89 | NS | NS |
| **Surgical_Technique_Right trisectionectomy + vascular resection** | Chi2 con permutación de Monte Carlo | 0,24 | NS | NS |
| **Segmente_1_Yes** | Chi2 test | 0,66 | NS | NS |
| **Pringle_maneuver_Yes** | Chi2 test | 0,27 | NS | NS |
| **RBC_Transfusion_Yes** | Chi2 test | 0,66 | NS | NS |
| **Total_number_of_lymph_nodes_resected_ge6** | Chi2 test | 0,61 | NS | NS |
| **Total_number_of_Positive_lymph_nodes_resected_ge6** | Chi2 con permutación de Monte Carlo | 0,24 | NS | NS |
| **Node_stage__pN__N1** | Chi2 test | 0,013 | Benjamini-Hochberg (FDR) | 1_a_3_years-<_1_year (pvalue: 0.22) // >_3_years-<_1_year (pvalue: 0.021) // >_3_years-1_a_3_years (pvalue: 0.23) |
| **HistologicGrade_G1** | Chi2 test | 0,095 | NS | NS |
| **HistologicGrade_G2** | Chi2 test | 0,16 | NS | NS |
| **HistologicGrade_G3** | Chi2 con permutación de Monte Carlo | 1 | NS | NS |
| **Distal_duodenal_margin_status_Negative** | Chi2 test | 0,25 | NS | NS |
| **Distal_duodenal_margin_status_Positive** | Chi2 con permutación de Monte Carlo | 1 | NS | NS |
| **Distal_duodenal_margin_status_ Positive with invasive cancer** | Chi2 con permutación de Monte Carlo | 0,031 | Benjamini-Hochberg (FDR) | 1_a_3_years-<_1_year (pvalue: 0.077) // >_3_years-<_1_year (pvalue: 1) // >_3_years-1_a_3_years (pvalue: 0.077) |
| **Distal_duodenal_margin_status_ Positive with Carcinoma in situ** | Chi2 con permutación de Monte Carlo | 0,74 | NS | NS |
| **Surgical_margin_status_R1** | Chi2 test | 0,51 | NS | NS |
| **Lymphatic_vessel_invasion_Yes** | Chi2 test | 0,68 | NS | NS |
| **Perineural_invasion_Yes** | Chi2 test | 0,067 | NS | NS |
| **Vascular_invasion_Yes** | Chi2 test | 0,45 | NS | NS |

| | | | | |
|---|---|---|---|---|
| **Adjuvant_chemotherapy_Yes** | Chi2 test | 0,27 | NS | NS |
| **Adjuvant_radiotherapy_Yes** | Chi2 test | 0,46 | NS | NS |
| **PO_Blood_Transfusion_Yes** | Chi2 test | 0,93 | NS | NS |
| **Hospital_readmission_Yes** | Chi2 test | 0,12 | NS | NS |
| **Grading_for_PHLF_ISGLS_C** | Chi2 con permutación de Monte Carlo | 0,62 | NS | NS |
| **Grading_for_PHLF_ISGLS_B** | Chi2 con permutación de Monte Carlo | 0,78 | NS | NS |
| **Grading_for_PHLF_ISGLS_A** | Chi2 con permutación de Monte Carlo | 0,83 | NS | NS |
| **Recurrence_status_Yes** | Chi2 test | 0,022 | Benjamini-Hochberg (FDR) | 1_a_3_years-<_1_year (pvalue: 0.034) // >_3_years-<_1_year (pvalue: 0.23) // >_3_years-1_a_3_years (pvalue: 0.23) |

#### 4.2. *Supplementary Table 2*

| Variable | Test | p.value |
|---|---|---|
| **Age** | Mann-Whitney test | 0,057 |
| **BMI** | Mann-Whitney test | 0,5 |
| **Pre_Bilirrubin** | Mann-Whitney test | 0,53 |
| **Pre_Ca19_9** | Mann-Whitney test | 0,77 |
| **Primary_tumor** | Mann-Whitney test | 0,21 |
| **Staging_AJCC_8th_ed_** | Mann-Whitney test | 0,0021 |
| **ICU_days** | Mann-Whitney test | 0,07 |
| **Hospital_total_days** | Mann-Whitney test | 0,031 |
| **Days_follow_up_or_death** | Mann-Whitney test | 0,72 |
| **Vascular_resection** | Fisher test | 0,74 |
| **Primary_tumor_dichotomic** | Chi2 test | 0,019 |
| **Staging_AJCC_8th_ed__dichotomic** | Chi2 test | 0,016 |
| **Clavien** | Mann-Whitney test | 0,42 |
| **Clavien_dichotomic** | Chi2 test | 0,97 |
| **Patient_status_cut** | Mann-Whitney test | 0,33 |
| **Gender_F** | Chi2 test | 0,52 |
| **Comorbidities_Yes** | Chi2 test | 0,3 |
| **endoscopic_biopsy_sample_Yes** | Chi2 test | 0,97 |
| **Portal_Vein_Embolization_Yes** | Chi2 test | 0,2 |
| **Preoperative_biliary_drainage_CatB_EBSoENBD** | Chi2 test | 1 |
| **Preoperative_biliary_drainage_CatC_PTCD** | Chi2 test | 0,085 |
| **Surgical_Technique_Bile duct resection** | Chi2 test | 0,88 |
| **Surgical_Technique_Bile duct resection + 4b-5 segmentectomy** | Fisher test | 1 |
| **Surgical_Technique_Bile duct resection + vascular resection** | Fisher test | 1 |
| **Surgical_Technique_Left hepatectomy** | Chi2 test | 0,95 |
| **Surgical_Technique_Left hepatectomy + vascular resection** | Fisher test | 0,65 |
| **Surgical_Technique_Left trisectionectomy** | Fisher test | 1 |
| **Surgical_Technique_Mesohepatectomy** | Fisher test | 0,39 |
| **Surgical_Technique_Right hepatectomy** | Chi2 test | 0,65 |
| **Surgical_Technique_Right hepatectomy + vascular resection** | Fisher test | 0,52 |
| **Surgical_Technique_Right trisectionectomy** | Chi2 test | 0,96 |
| **Surgical_Technique_Right trisectionectomy + vascular resection** | Fisher test | 0,39 |
| **Segmente_1_Yes** | Chi2 test | 0,66 |
| **Pringle_maneuver_Yes** | Chi2 test | 1 |
| **RBC_Transfusion_Yes** | Chi2 test | 0,22 |
| **Total_number_of_lymph_nodes_resected_ge6** | Chi2 test | 0,22 |
| **Total_number_of_Positive_lymph_nodes_resected_ge6** | Fisher test | 1 |
| **Node_stage__pN__N1** | Chi2 test | 0,016 |
| **HistologicGrade_G1** | Chi2 test | 0,91 |
| **HistologicGrade_G2** | Chi2 test | 0,37 |
| **HistologicGrade_G3** | Fisher test | 0,2 |

| | | |
|---|---|---|
| **Distal_duodenal_margin_status_Negative** | Chi2 test | 0,082 |
| **Distal_duodenal_margin_status_Positive** | Fisher test | 1 |
| **Distal_duodenal_margin_status_ Positive with invasive cancer** | Chi2 test | 0,27 |
| **Distal_duodenal_margin_status_ Positive with Carcinoma in situ** | Fisher test | 1 |
| **Surgical_margin_status_R1** | Chi2 test | 0,11 |
| **Lymphatic_vessel_invasion_Yes** | Chi2 test | 0,089 |
| **Perineural_invasion_Yes** | Chi2 test | 0,011 |
| **Vascular_invasion_Yes** | Chi2 test | 0,067 |
| **Adjuvant_chemotherapy_Yes** | Chi2 test | 0,0085 |
| **Adjuvant_radiotherapy_Yes** | Chi2 test | 0,96 |
| **PO_Blood_Transfusion_Yes** | Chi2 test | 0,053 |
| **Hospital_readmission_Yes** | Chi2 test | 1 |
| **Grading_for_PHLF_ISGLS_C** | Fisher test | 0,28 |
| **Grading_for_PHLF_ISGLS_B** | Fisher test | 0,74 |
| **Grading_for_PHLF_ISGLS_A** | Fisher test | 0,31 |

#### 4.3. *Supplementary Table 3*

| Variable | Test | p.value |
|---|---|---|
| **Age** | Kruskal-Wallis test | 0,24 |
| **BMI** | Kruskal-Wallis test | 0,48 |
| **Pre_Bilirrubin** | Kruskal-Wallis test | 0,031 |
| **Pre_Ca19_9** | Kruskal-Wallis test | 0,46 |
| **Primary_tumor** | Kruskal-Wallis Test | 0,00003 |
| **Staging_AJCC_8th_ed_** | Kruskal-Wallis Test | 7,2E-14 |
| **ICU_days** | Kruskal-Wallis test | 0,35 |
| **Hospital_total_days** | Kruskal-Wallis test | 0,34 |
| **Days_follow_up_or_death** | Kruskal-Wallis test | 0,028 |
| **Vascular_resection** | Chi2 con permutación de Monte Carlo | 0,78 |
| **Primary_tumor_dicotomica** | Chi2 con permutación de Monte Carlo | 0,0015 |
| **Staging_AJCC_8th_ed__dicotomica** | Chi2 test | 5,6E-13 |
| **Days_recurrence** | Kruskal-Wallis test | 0,062 |
| **Clavien** | Kruskal-Wallis Test | 0,8 |
| **Clavien_dicotomica** | Chi2 test | 0,56 |
| **Gender_F** | Chi2 test | 0,097 |
| **Comorbidities_Yes** | Chi2 test | 0,0007 |
| **endoscopic_biopsy_sample_Yes** | Chi2 test | 0,13 |
| **Portal_Vein_Embolization_Yes** | Chi2 test | 0,24 |
| **Preoperative_biliary_drainage_CatB_EBSoENBD** | Chi2 test | 0,36 |
| **Preoperative_biliary_drainage_CatC_PTCD** | Chi2 test | 0,32 |
| **Surgical_Technique_Bile duct resection** | Chi2 test | 1,3E-09 |
| **Surgical_Technique_Bile duct resection + 4b-5 segmentectomy** | Chi2 con permutación de Monte Carlo | 0,35 |
| **Surgical_Technique_Bile duct resection + vascular resection** | Chi2 con permutación de Monte Carlo | 0,69 |
| **Surgical_Technique_Left hepatectomy** | Chi2 test | 0,0012 |
| **Surgical_Technique_Left hepatectomy + vascular resection** | Chi2 con permutación de Monte Carlo | 0,73 |
| **Surgical_Technique_Left trisectionectomy** | Chi2 con permutación de Monte Carlo | 0,33 |
| **Surgical_Technique_Mesohepatectomy** | Chi2 con permutación de Monte Carlo | 0,6 |
| **Surgical_Technique_Right hepatectomy** | Chi2 test | 0,009 |
| **Surgical_Technique_Right hepatectomy + vascular resection** | Chi2 con permutación de Monte Carlo | 0,69 |
| **Surgical_Technique_Right trisectionectomy** | Chi2 con permutación de Monte Carlo | 0,17 |
| **Surgical_Technique_Right trisectionectomy + vascular resection** | Chi2 con permutación de Monte Carlo | 0,6 |
| **Segmente_1_Yes** | Chi2 test | 1,1E-18 |
| **Pringle_maneuver_Yes** | Chi2 test | 0,000047 |
| **RBC_Transfusion_Yes** | Chi2 test | 0,00027 |
| **Total_number_of_lymph_nodes_resected_ge6** | Chi2 test | 0,61 |
| **Total_number_of_Positive_lymph_nodes_resected_ge6** | Chi2 con permutación de Monte Carlo | 0,22 |

| | | |
|---|---|---|
| **Node_stage__pN__N1** | Chi2 test | 3E-15 |
| **HistologicGrade_G1** | Chi2 test | 0,011 |
| **HistologicGrade_G2** | Chi2 test | 0,29 |
| **HistologicGrade_G3** | Chi2 con permutación de Monte Carlo | 0,025 |
| **Distal_duodenal_margin_status_ Negative** | Chi2 test | 0,00008 |
| **Distal_duodenal_margin_status_P ositive** | Chi2 con permutación de Monte Carlo | 1 |
| **Distal_duodenal_margin_status_ Positive with invasive cancer** | Chi2 con permutación de Monte Carlo | 0,00006 |
| **Distal_duodenal_margin_status_ Positive with Carcinoma in situ** | Chi2 con permutación de Monte Carlo | 0,84 |
| **Surgical_margin_status_R1** | Chi2 test | 0,00023 |
| **Lymphatic_vessel_invasion_Yes** | Chi2 test | 6,8E-09 |
| **Perineural_invasion_Yes** | Chi2 test | 0,035 |
| **Vascular_invasion_Yes** | Chi2 test | 4,6E-10 |
| **Adjuvant_chemotherapy_Yes** | Chi2 test | 0,019 |
| **Adjuvant_radiotherapy_Yes** | Chi2 test | 0,073 |
| **PO_Blood_Transfusion_Yes** | Chi2 test | 0,18 |
| **Hospital_readmission_Yes** | Chi2 test | 0,72 |
| **Grading_for_PHLF_ISGLS_C** | Chi2 con permutación de Monte Carlo | 0,18 |
| **Grading_for_PHLF_ISGLS_B** | Chi2 con permutación de Monte Carlo | 0,1 |
| **Grading_for_PHLF_ISGLS_A** | Chi2 con permutación de Monte Carlo | 0,91 |
| **Patient_status_Exitus** | Chi2 test | 0,19 |
| **Recurrence_status_Yes** | Chi2 test | 0,11 |
| **Months_recurrence** | Kruskal-Wallis test | 0,062 |
| **Months_follow_up_or_death** | Kruskal-Wallis test | 0,028 |

#### *4.4. Supplementary Table 4*

| Variable | Cluster 2-Cluster 1 | Cluster 3-Cluster 1 | Cluster 3-Cluster 2 |
|---|---|---|---|
| Pre_Bilirrubin | NS | * | NS |
| Primary_tumor | NS | ** | *** |
| Staging_AJCC_8th_ed_ | NS | *** | *** |
| Days_follow_up_or_death | NS | * | NS |
| Primary_tumor_dicotomica | * | NS | * |
| Staging_AJCC_8th_ed__dicotomica | NS | *** | *** |
| Comorbidities_Yes | ** | NS | NS |
| Surgical_Technique_Bile duct resection | *** | NS | *** |
| Surgical_Technique_Left hepatectomy | ** | NS | NS |
| Surgical_Technique_Right hepatectomy | NS | * | NS |
| Segmente_1_Yes | *** | *** | *** |
| Pringle_maneuver_Yes | *** | NS | ** |
| RBC_Transfusion_Yes | *** | NS | ** |
| Node_stage__pN__N1 | NS | *** | *** |
| HistologicGrade_G1 | NS | * | NS |
| HistologicGrade_G3 | NS | NS | NS |
| Distal_duodenal_margin_status_Negative | NS | *** | ** |
| Distal_duodenal_margin_status_ Positive with invasive cancer | NS | ** | ** |
| Surgical_margin_status_R1 | NS | *** | * |
| Lymphatic_vessel_invasion_Yes | NS | *** | *** |
| Perineural_invasion_Yes | NS | NS | NS |
| Vascular_invasion_Yes | NS | *** | *** |
| Adjuvant_chemotherapy_Yes | NS | * | NS |
| Months_follow_up_or_death | NS | * | NS |

#### 4.5. *Supplementary Table 5*

| Variable | Test | p.value |
|---|---|---|
| **Age** | Anova test | 0,14 |
| **BMI** | Kruskal-Wallis test | 0,43 |
| **Pre_Bilirrubin** | Kruskal-Wallis test | 0,56 |
| **Pre_Ca19_9** | Kruskal-Wallis test | 0,96 |
| **Primary_tumor** | Kruskal-Wallis Test | 0,16 |
| **Staging_AJCC_8th_ed_** | Kruskal-Wallis Test | 0,0006 |
| **ICU_days** | Kruskal-Wallis test | 0,058 |
| **Hospital_total_days** | Kruskal-Wallis test | 0,91 |
| **Vascular_resection** | Chi2 con permutación de Monte Carlo | 0,55 |
| **Primary_tumor_dicotomica** | Chi2 con permutación de Monte Carlo | 0,18 |
| **Staging_AJCC_8th_ed__dicotomica** | Chi2 con permutación de Monte Carlo | 0,0024 |
| **Clavien** | Kruskal-Wallis Test | 0,42 |
| **Clavien_dicotomica** | Chi2 con permutación de Monte Carlo | 0,68 |
| **Patient_status_cut** | Kruskal-Wallis Test | 0,21 |
| **Gender_F** | Chi2 con permutación de Monte Carlo | 0,77 |
| **Comorbidities_Yes** | Chi2 con permutación de Monte Carlo | 0,35 |
| **endoscopic_biopsy_sample_Yes** | Chi2 con permutación de Monte Carlo | 0,37 |
| **Portal_Vein_Embolization_Yes** | Chi2 con permutación de Monte Carlo | 0,72 |
| **Preoperative_biliary_drainage_CatB_EBSoENBD** | Chi2 con permutación de Monte Carlo | 0,35 |
| **Preoperative_biliary_drainage_CatC_PTCD** | Chi2 con permutación de Monte Carlo | 0,29 |
| **Surgical_Technique_Bile duct resection** | Chi2 con permutación de Monte Carlo | 0,016 |
| **Surgical_Technique_Bile duct resection + 4b-5 segmentectomy** | Chi2 con permutación de Monte Carlo | 1 |
| **Surgical_Technique_Bile duct resection + vascular resection** | Chi2 con permutación de Monte Carlo | 0,56 |
| **Surgical_Technique_Left hepatectomy** | Chi2 con permutación de Monte Carlo | 0,14 |
| **Surgical_Technique_Left hepatectomy + vascular resection** | Chi2 con permutación de Monte Carlo | 1 |
| **Surgical_Technique_Left trisectionectomy** | Chi2 con permutación de Monte Carlo | 1 |
| **Surgical_Technique_Mesohepatectomy** | Chi2 con permutación de Monte Carlo | 1 |
| **Surgical_Technique_Right hepatectomy** | Chi2 con permutación de Monte Carlo | 0,55 |
| **Surgical_Technique_Right hepatectomy + vascular resection** | Chi2 con permutación de Monte Carlo | 1 |
| **Surgical_Technique_Right trisectionectomy** | Chi2 con permutación de Monte Carlo | 0,83 |
| **Surgical_Technique_Right trisectionectomy + vascular resection** | Chi2 con permutación de Monte Carlo | 1 |
| **Segmente_1_Yes** | Chi2 con permutación de Monte Carlo | 0,00015 |
| **Pringle_maneuver_Yes** | Chi2 con permutación de Monte Carlo | 0,55 |
| **RBC_Transfusion_Yes** | Chi2 con permutación de Monte Carlo | 0,18 |
| **Total_number_of_lymph_nodes_resected_ge6** | Chi2 con permutación de Monte Carlo | 0,41 |

| | | |
|---|---|---|
| **Total_number_of_Positive_lymph_nodes_resected_ge6** | Chi2 con permutación de Monte Carlo | 0,56 |
| **Node_stage__pN__N1** | Chi2 con permutación de Monte Carlo | 0,00004 |
| **HistologicGrade_G1** | Chi2 con permutación de Monte Carlo | 0,049 |
| **HistologicGrade_G2** | Chi2 con permutación de Monte Carlo | 0,25 |
| **HistologicGrade_G3** | Chi2 con permutación de Monte Carlo | 0,57 |
| **Distal_duodenal_margin_status_Negative** | Chi2 con permutación de Monte Carlo | 0,13 |
| **Distal_duodenal_margin_status_Positive** | Chi2 con permutación de Monte Carlo | 1 |
| **Distal_duodenal_margin_status_ Positive with invasive cancer** | Chi2 con permutación de Monte Carlo | 0,025 |
| **Distal_duodenal_margin_status_ Positive with Carcinoma in situ** | Chi2 con permutación de Monte Carlo | 1 |
| **Surgical_margin_status_R1** | Chi2 con permutación de Monte Carlo | 0,45 |
| **Lymphatic_vessel_invasion_Yes** | Chi2 con permutación de Monte Carlo | 0,0023 |
| **Perineural_invasion_Yes** | Chi2 con permutación de Monte Carlo | 0,62 |
| **Vascular_invasion_Yes** | Chi2 con permutación de Monte Carlo | 0,00003 |
| **Adjuvant_chemotherapy_Yes** | Chi2 con permutación de Monte Carlo | 0,21 |
| **Adjuvant_radiotherapy_Yes** | Chi2 con permutación de Monte Carlo | 0,56 |
| **PO_Blood_Transfusion_Yes** | Chi2 con permutación de Monte Carlo | 0,0092 |
| **Hospital_readmission_Yes** | Chi2 con permutación de Monte Carlo | 0,71 |
| **Grading_for_PHLF_ISGLS_C** | Chi2 con permutación de Monte Carlo | 0,56 |
| **Grading_for_PHLF_ISGLS_B** | Chi2 con permutación de Monte Carlo | 0,017 |
| **Grading_for_PHLF_ISGLS_A** | Chi2 con permutación de Monte Carlo | 1 |
| **Patient_status_Exitus** | Chi2 con permutación de Monte Carlo | 1 |
| **Recurrence_status_Yes** | Chi2 con permutación de Monte Carlo | 1 |

#### 4.6. ***Supplementary Table 6***

| Variable | C2 <8months-C1 <8months | C3 <8months-C1 <8months | C3 <8months-C2 <8months |
|---|---|---|---|
| **Staging_AJCC_8th_ed_** | NS | * | *** |
| **Staging_AJCC_8th_ed__dicotomica** | NS | * | * |
| **Surgical_Technique_Bile duct resection** | NS | NS | NS |
| **Segmente_1_Yes** | ** | NS | * |
| **Node_stage__pN__N1** | NS | * | *** |
| **HistologicGrade_G1** | NS | NS | NS |
| **Distal_duodenal_margin_status_ Positive with invasive cancer** | | NS | NS |
| **Lymphatic_vessel_invasion_Yes** | NS | NS | * |
| **Vascular_invasion_Yes** | NS | * | ** |
| **PO_Blood_Transfusion_Yes** | NS | NS | NS |
| **Grading_for_PHLF_ISGLS_B** | NS | NS | |

#### 4.7. Supplementary Table 7

| Variable | Beta-value | Percentage explained |
|---|---|---|
| chr1_27023514_T_A | 1,14 | 4,14 |
| chr19_1220414_C_T | 1,06 | 3,84 |
| chr1_27022907_G_A | 0,85 | 3,08 |
| Gender_F | 0,79 | 2,87 |
| chr4_55979558_C_T | 0,78 | 2,83 |
| PO_Blood_Transfusion_Yes | 0,71 | 2,57 |
| chr4_55593513_T_C | 0,59 | 2,13 |
| chr1_27023523_A_G | 0,56 | 2,01 |
| Surgical_Technique_Right_trisectionectomy | 0,50 | 1,80 |
| chr1_27023347_G_A | 0,49 | 1,78 |
| Comorbidities_Yes | 0,49 | 1,77 |
| Pringle_maneuver_Yes | 0,47 | 1,71 |
| Staging_AJCC_8th_ed_ | 0,46 | 1,66 |
| HistologicGrade_G1 | 0,45 | 1,64 |
| Grading_for_PHLF_ISGLS_B | 0,41 | 1,48 |
| Vascular_invasion_Yes | 0,40 | 1,46 |
| endoscopic_biopsy_sample_Yes | 0,39 | 1,43 |
| chr13_28610183_A_G | 0,39 | 1,41 |
| Hospital_readmission_Yes | 0,38 | 1,39 |
| chr22_24176287_G_A | 0,36 | 1,32 |
| Portal_Vein_Embolization_Yes | 0,36 | 1,29 |
| Adjuvant_radiotherapy_Yes | 0,34 | 1,23 |
| chr18_48586344_C_T | 0,32 | 1,17 |
| chr9_22006175_G_A | 0,31 | 1,13 |
| chr10_43615633_C_G | 0,30 | 1,09 |
| Grading_for_PHLF_ISGLS_A | 0,20 | 0,72 |
| BMI | 0,18 | 0,66 |
| Lymphatic_vessel_invasion_Yes | 0,16 | 0,59 |
| chr7_55248926_T_C | 0,15 | 0,53 |
| Surgical_Technique_Left_hepatectomy | 0,14 | 0,49 |
| chr4_55593464_A_C | 0,14 | 0,49 |
| Preoperative_biliary_drainage_CatC_PTCD | 0,12 | 0,42 |
| Pre_Bilirrubin | 0,11 | 0,40 |
| chr19_1220402_G_A | 0,10 | 0,38 |
| chr1_27057621_A_C | 0,10 | 0,37 |
| Surgical_margin_status_R1 | 0,08 | 0,28 |
| chr3_178927410_A_G | 0,07 | 0,27 |
| chr12_121431300_C_T | 0,07 | 0,24 |
| chr7_128846328_G_C | 0,06 | 0,23 |
| Surgical_Technique_Right_hepatectomy | 0,06 | 0,21 |
| chr13_28602256_C_T | 0,05 | 0,19 |

| | | |
|---|---|---|
| **chr3_178938747_A_T** | 0,04 | 0,16 |
| **chr1_27100243_C_T** | 0,03 | 0,12 |
| **Age** | 0,03 | 0,11 |
| **Adjuvant_chemotherapy_Yes** | 0,01 | 0,05 |
| **chr4_55972974_T_A** | 0,00 | 0,00 |
| **Preoperative_biliary_drainage_CatB_EBSo_ENBD** | 0,00 | 0,00 |
| **RBC_Transfusion_Yes** | -0,01 | 0,05 |
| **Pre_Ca19_9** | -0,03 | 0,11 |
| **chr17_7578210_T_C** | -0,05 | 0,20 |
| **HistologicGrade_G3** | -0,08 | 0,28 |
| **chr13_28592546_T_C** | -0,09 | 0,34 |
| **Preoperative_biliary_drainage_CatA_Nodrainage** | -0,12 | 0,42 |
| **Distal_duodenal_margin_status_Positive_with_invasive_cancer** | -0,12 | 0,44 |
| **chr7_128846075_C_T** | -0,13 | 0,49 |
| **chr5_112175770_G_A** | -0,14 | 0,49 |
| **chr14_105246407_G_A** | -0,15 | 0,53 |
| **Perineural_invasion_Yes** | -0,16 | 0,57 |
| **chr7_55248862_C_T** | -0,17 | 0,60 |
| **Primary_tumor** | -0,18 | 0,64 |
| **chr9_21971018_G_A** | -0,18 | 0,66 |
| **chr9_21971024_G_A** | -0,20 | 0,74 |
| **Node_stage__pN__N1** | -0,21 | 0,77 |
| **chr13_28602292_T_C** | -0,24 | 0,86 |
| **chr2_212589800_C_T** | -0,25 | 0,89 |
| **chr3_178917005_A_G** | -0,25 | 0,90 |
| **Surgical_Technique_Bile_duct_resection** | -0,29 | 1,07 |
| **chr7_55259550_C_T** | -0,30 | 1,08 |
| **chr7_55249063_G_A** | -0,30 | 1,08 |
| **chr4_55946081_A_G** | -0,32 | 1,14 |
| **chr9_21971019_C_T** | -0,34 | 1,25 |
| **Distal_duodenal_margin_status_Negative** | -0,35 | 1,28 |
| **HistologicGrade_G2** | -0,37 | 1,36 |
| **chr1_27023348_C_T** | -0,46 | 1,67 |
| **Vascular_resection** | -0,46 | 1,67 |
| **chr12_56494991_G_A** | -0,54 | 1,95 |
| **chr9_21971021_G_A** | -0,55 | 1,98 |
| **Segmente_1_Yes** | -0,55 | 1,99 |
| **chr4_55602765_G_C** | -0,59 | 2,13 |
| **chr14_105241378_C_T** | -0,60 | 2,16 |
| **chr12_56494998_A_T** | -0,61 | 2,22 |
| **Total_number_of_lymph_nodes_resected_>=_6** | -0,83 | 3,02 |
| **chr1_27023345_G_A** | -0,96 | 3,47 |
| **chr17_7573897_T_A** | -1,20 | 4,33 |
